# Clinical-grade autonomous cytopathology via whole-slide edge tomography

**DOI:** 10.1101/2025.06.26.25330376

**Authors:** Nao Nitta, Yuko Sugiyama, Takeaki Sugimura, Takahiko Ito, Koichi Ikebata, Hitoshi Abe, Shuhei Ishii, Nagisa Hosoya, Raihan Ull Islam, Aditya Jain, Meisam Hasani, Joseph Zonghi, Peter Koh, Yukihito Mase, Yingdong Luo, Tianben Ding, Fernando C. Schmitt, Robert Y. Osamura, Keisuke Goda, Tomohiro Chiba

## Abstract

Cytopathology plays a central role in the early detection of cancers such as cervical, lung, and bladder cancer due to its speed, simplicity, and minimally invasive nature. However, its effectiveness is limited by variability in diagnostic accuracy stemming from subjective visual interpretation. Although many AI-powered systems have been proposed to improve consistency, none have achieved fully autonomous, clinical-grade performance. Existing approaches serve as assistive tools and still rely on human oversight for interpretation and decision-making. Here we present a clinical-grade autonomous cytopathology pipeline that combines high-resolution, real-time optical whole-slide tomography with edge computing to deliver end-to-end automation. The system achieves practical performance in imaging speed, quality, and data volume, with localized data compression enabling streamlined storage and accelerated AI-driven analysis. In addition to supporting cell-level classification, the platform enables flow cytometry-like, population-wide morphological profiling for comprehensive interpretation of cellular distributions and patterns. A vision transformer achieved area-under-receiver-operating-characteristic-curve values exceeding 0.99 for detecting LSIL, HSIL, and adenocarcinoma. In a clinical cohort of cervical liquid-based cytology samples from 318 donors, LSIL counts strongly correlated with HPV positivity, while HSIL counts scaled with diagnostic severity. The system enables autonomous triage cytology, offering a foundation for routine, scalable, and objective diagnostics.

## INTRODUCTION

Cytopathology, often abbreviated as cytology, lies at the core of early cancer detection, particularly for cervical, lung, and bladder malignancies, due to its minimally invasive nature, affordability, and diagnostic utility^1–7^. In cytological examinations, cytologists employ optical microscopes to identify and assess potential abnormalities in the architecture of individual cells or cell clusters based on their three-dimensional (3D) structure, morphology, and relationships with neighboring cells. These assessments are conducted on glass slides that typically contain approximately 10,000 to 1,000,000 cells per slide. The Pap smear, the most common cytological test, specifically targets early detection of pre-cancerous or abnormal cervical cells^8,9^. While traditionally performed by directly smearing cells onto slides, Pap testing is increasingly conducted using liquid-based cytology, a method that has significantly contributed to reducing cervical cancer incidence and mortality through improved sample quality and diagnostic accuracy^4,5^.

Despite its benefits, cytology suffers from variability in diagnostic sensitivity and specificity, largely due to subjectivity in visual interpretation^10,11^. Differences in training and experience levels can lead to varying interpretations of cytological slides, while cognitive biases such as confirmation and anchoring bias can skew diagnoses^12–16^. Inconsistent adherence to standardized guidelines further exacerbates these issues^17,18^. Factors such as fatigue and high workload also impact diagnostic accuracy, contributing to frequent misdiagnoses and overlooked cases^16^. The CervicalCheck cancer scandal in Ireland^19–21^ illustrates the potential clinical consequences of subjective cytology and highlights the pressing need for objective, reproducible diagnostics. Manual screening is further challenged by the low frequency of abnormal cells, presence of preparation artefacts, and overlapping cellular structures, all of which hinder early disease detection^22,23^. The incorporation of advanced technologies, uniform diagnostic criteria, and consistent adherence to guidelines could greatly reduce misdiagnoses and ensure more reliable, objective, and equitable medical assessments, enabling timely and appropriate clinical interventions.

Artificial intelligence (AI) has been widely explored to support cytological interpretation, with various tools developed for cell classification and region detection^15,22,24–26^. However, most of these approaches rely on two-dimensional (2D) images and focus primarily on identifying isolated representative or suspicious cells. This reflects a more fundamental limitation: the absence of frameworks capable of large-scale, quantitative analysis across thousands of spatially dispersed cells. Although 3D imaging offers superior spatial detail, it also introduces substantial challenges in image acquisition, processing, and storage. Compared to histopathology and radiology^27–32^, cytology generates far greater data volumes due to the requirement for depth-resolved imaging of morphologically diverse, non-cohesive cells. These demands on storage capacity, computational power, and data transfer bandwidth have limited the scalability of AI models. As a result, most cytology AI applications remain assistive, still relying on human interpretation and decision-making, rather than functioning autonomously^15,22,24–26^. To date, no AI-based system has demonstrated clinical-grade performance in autonomous cytological diagnosis.

To overcome these limitations, we present a real-time, clinically validated, autonomous cytology platform that integrates high-speed, high-resolution optical whole-slide tomography with edge computing – a distributed computing architecture that processes data closer to its source^33–36^. This method enables the acquisition and local compression of high-quality 3D whole-slide images (140 or 393 gigavoxels per slide at lateral and axial resolutions of 220 nm and 1 μm, respectively) before storage, significantly speeding up training, validation, and deployment without compromising image quality or AI performance. This approach effectively digitizes thick cytology samples containing abnormal cell clusters – a long-standing technical challenge in cytology^37–39^. Notably, this high-quality 3D digitization enhances AI performance without requiring a large dataset from numerous patients for training; approximately 1,000 3D original images per class are sufficient for effective AI training.

A key innovation of our platform is the introduction of the cluster of morphological differentiation (CMD) – an image-derived analogue to the cluster of differentiation (CD) used in immunophenotyping^40,41^. The CMD enables a flow cytometry-like analytical framework based on morphological features extracted from imaging data, rather than fluorescence markers. Unlike earlier AI-based cytology tools^15,19–22,24–26^, the CMD allows cytologists to intuitively visualize and explore entire cell populations using tools such as scatter plots, hierarchical gating, and dimensionality reduction. These capabilities facilitate not only enhanced interpretability and error detection, but also the discovery of previously unrecognized cellular phenotypes. Together, these advances establish a scalable, real-time cytology pipeline with clinical-grade autonomy. By transforming cytology into a quantitative, population-scale discipline, the platform lays the foundation for a new diagnostic paradigm that is objective, reproducible, and discovery-driven.

## RESULTS

### Whole-slide edge tomography

As illustrated in Figure 1, our edge computer-integrated optical whole-slide tomograph, referred to as the whole-slide edge tomograph, comprises a light-emitting diode (LED) as a light source, an XY translation stage, a Z translation stage integrated with imaging optics and a CMOS image sensor, and an edge computer equipped with an image sensor FPGA and a system-on-module (SOM) for real-time digital image processing (see Extended Data Figure 1 for details). This configuration enables the acquisition of 2D images across multiple depth layers and facilitates the construction, compression, and archiving of 3D images during slide scanning in the lateral (XY) and longitudinal (Z) directions. The CMOS image sensor captures high-resolution 2D bright-field images (4.5k x 4.5k pixels per imaging section) at a rate of up to 50 frames per second (fps), with 173 or 485 imaging sections per layer for SurePath (Becton Dickinson and Co.) or ThinPrep (Hologic) slides, respectively, and 40 layers per slide, yielding approximately 140 or 393 gigavoxels per slide, respectively. These 2D images are transmitted to the FPGA for initial image processing. The processed images are then sent to the GPU via a dual 4-lane MIPI interface, where additional tasks such as background correction, focus adjustment, 3D image construction, and 3D image compression are performed using the hardware encoder, a dedicated module in the SOM for real-time image compression. Similar to video compression, where redundancies within each frame (intra-frame) and between consecutive frames (inter-frame) reduce data size, our system exploits both intra-layer compression, by reducing spatial redundancy within individual 2D images, and inter-layer compression, by exploiting similarities between adjacent optical sections along the Z-axis. This strategy enables efficient compression of sectional 3D image stacks while preserving diagnostic content. The compressed images are stored in the high-efficiency video coding (HEVC) format, which supports both intra- and inter-prediction schemes and is well suited for volumetric image sequences. These sectional 3D images are then transmitted to a backend server, where they are decoded and stitched into a comprehensive 3D image of the entire slide, covering approximately 10,000 to 1,000,000 cells. This whole-slide 3D image can be viewed in real time by cytologists on a high-definition monitor, allowing interactive functions such as movement, magnification, and focus adjustments using a deep zoom image (DZI) viewer. Simultaneously, the 3D image undergoes AI-based population analysis. Nuclei in individual cells are detected using the high-performance object detection model YOLOX^42^, followed by cropping the best focused objects and classification with the vision transformer model MaxViT^43^. Additionally, population analysis is conducted using the CMD, with results provided to the cytologists. Both the 3D image and AI-generated analysis results are accessible in real time, supporting efficient diagnosis and evaluation.

**Figure 1.**
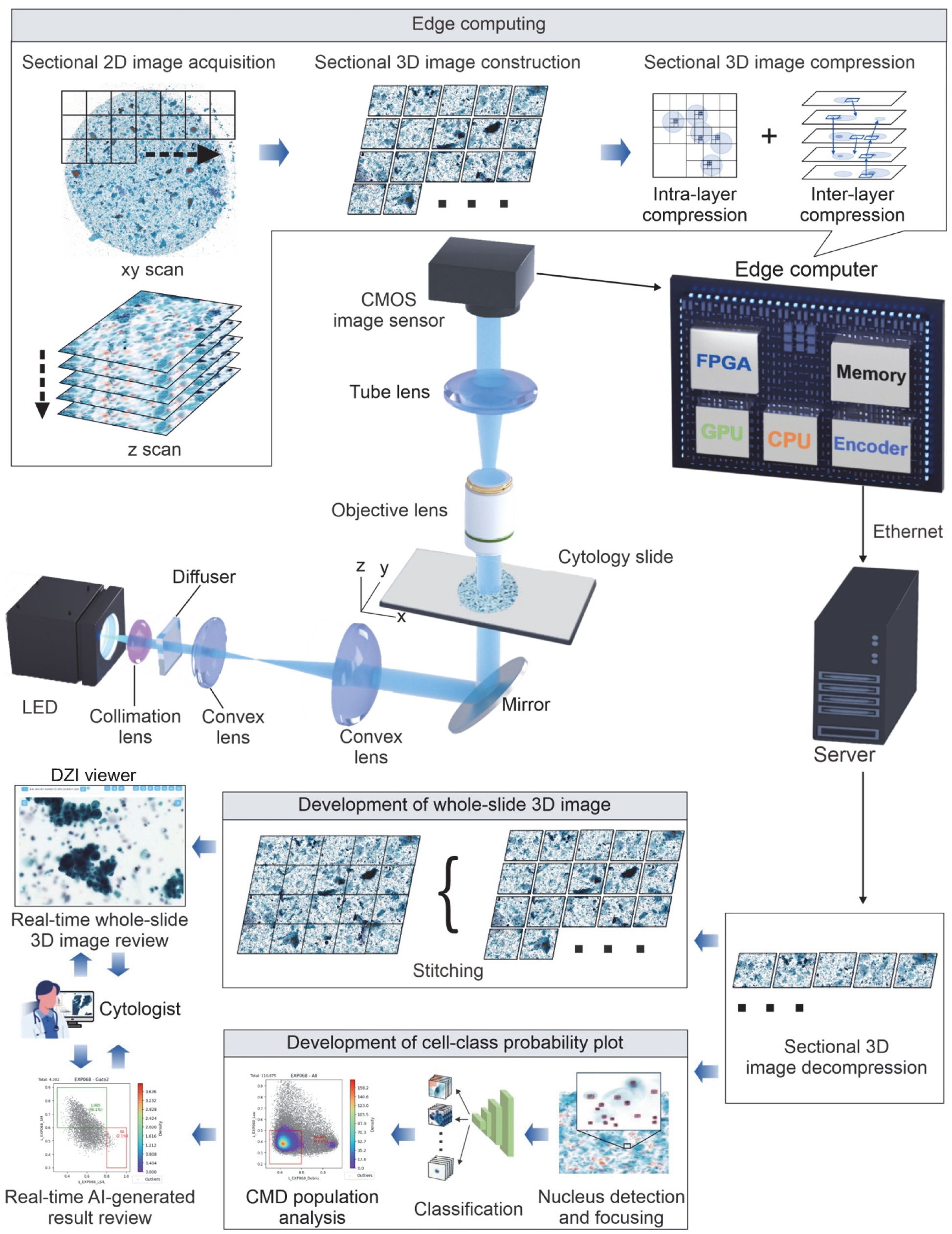
Whole-slide edge tomography. The edge computer facilitates the acquisition of 2D images across multiple depth layers and enables the construction, compression, and archiving of 3D images during slide scanning in both the lateral (XY) and longitudinal (Z) directions. The CMOS image sensor captures high-resolution 2D bright-field images (4.5k x 4.5k pixels per imaging section) at up to 50 frames per second, with 173 or 485 imaging sections per layer and 40 depth layers per slide, yielding approximately 140 or 393 gigavoxels per SurePath or ThinPrep slide, respectively. These 2D images are first transmitted to the FPGA within the edge computer for initial processing. Subsequently, they are sent to the GPU, where further tasks such as background correction, focus adjustment, 3D image construction, and compression are carried out. The processed sectional 3D images are then transmitted to a server, where the server’s GPU stitches them together into a comprehensive 3D representation of the entire slide, encompassing approximately 10,000 to 1,000,000 cells. This whole-slide 3D image can be viewed in real time by cytologists on a high-definition monitor, enabling interactive functions such as movement, magnification, and focus adjustments. Simultaneously, AI-based population analysis is performed on the 3D image. Population analysis is further refined using the CMD, and the results are provided to the cytologists. Both the 3D image and AI-generated analysis results are accessible in real time, enabling efficient diagnosis and evaluation.

### System performance

As shown in Figure 2, the whole-slide edge tomograph achieves high-quality imaging and efficient data compression. Figure 2a presents the image quality under different HEVC compression settings (high, medium, low) across three liquid-based cytology slides. For each condition, 300 tomographic images (3 slides x 10 imaging sections × 10 layers) were acquired, and peak signal-to-noise ratio (PSNR) distributions are shown as histograms. Figure 2b summarizes the corresponding file sizes of the 3D whole-slide images, demonstrating an inverse relationship between compression level and data size: approximately 1 GB, 500–800 MB, and 170 MB for high, medium, and low compression of a 10-layer SurePath slide, respectively. File size scaled linearly with the number of Z-layers. Figure 2c displays representative tomograms (2D cross-sections) of glandular, LSIL, HSIL, and adenocarcinoma cells across varying PSNR levels. The system resolves subcellular structures such as nucleoli and nuclear membranes with high fidelity at PSNR ≥ 40 dB, corresponding to medium-to-high compression. Figure 2d and Extended Data Figures 2a and 2b detail the imaging timeline across different Z-layer settings (10, 20, and 40 layers) for the first 8 imaging sections. The time logs break down the process into XY stage motion, image acquisition, reconstruction, and compression. These operations were pipelined, allowing computation and data encoding to proceed in parallel with mechanical movements, minimizing idle time. Figure 2e quantifies task latency per imaging section, showing that while XY motion time remained constant, acquisition, processing, and encoding times scaled proportionally with the number of Z-layers. Extended Data Figure 2e reports total imaging times per slide of approximately 3, 4.5, and 8 min for 10, 20, and 40 layers, respectively, with minimal influence from compression setting. Furthermore, Extended Data Figure 3b confirms real-time system responsiveness: even with on-the-fly HEVC decompression during viewing, most high-resolution tile requests were completed within 100 msec. These results collectively demonstrate the system’s predictable, high-performance operation across varying imaging conditions.

**Figure 2.**
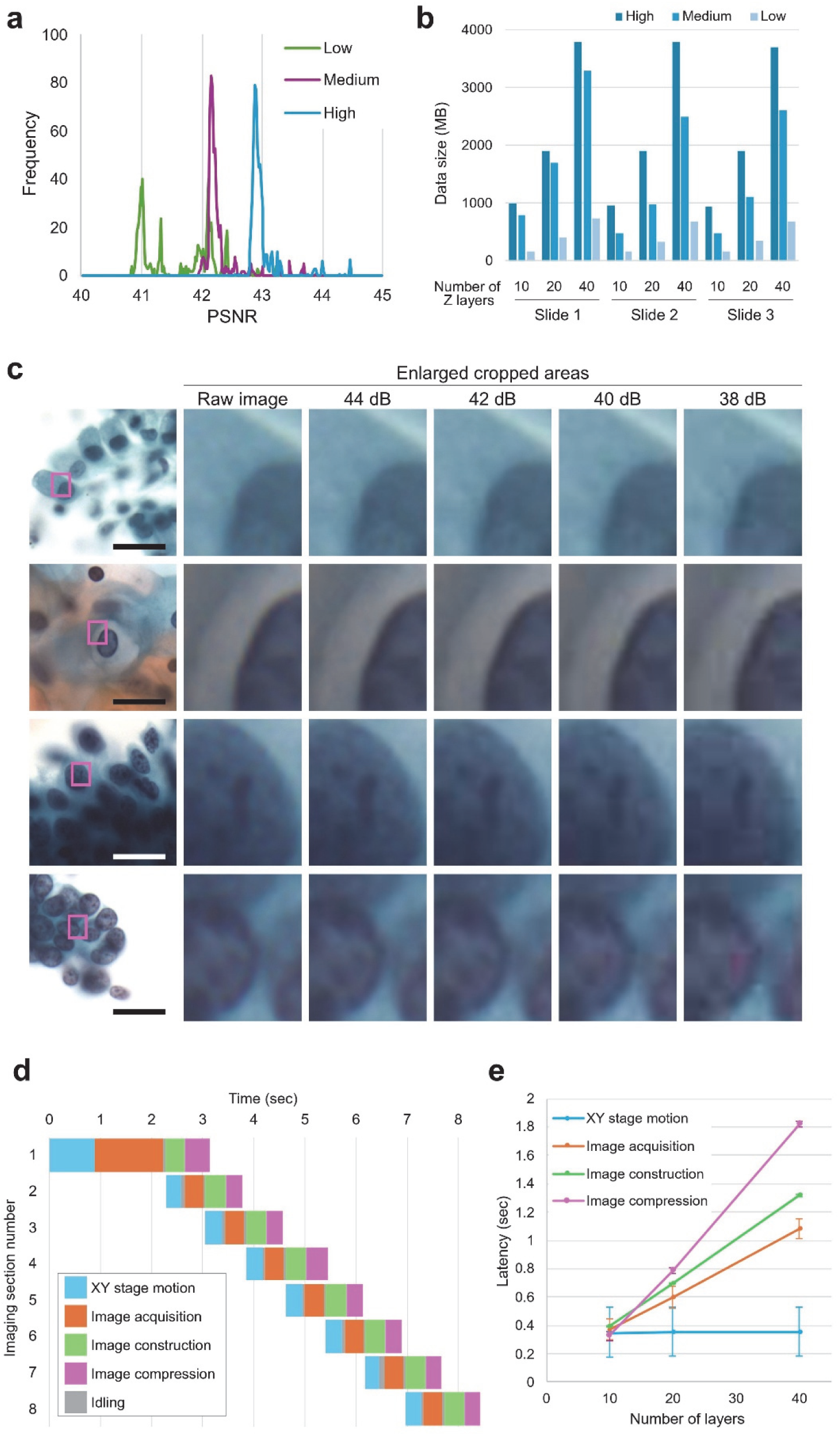
System performance. **a,** Histograms of PSNR values obtained from 300 tomographic images acquired from three liquid-based cytology slides under high, medium, and low HEVC compression settings (3 slides × 10 imaging sections × 10 Z-layers per slide). **b,** File sizes of whole-slide 3D image datasets under varying compression levels and Z-layer counts (10, 20, 40). Data size increases with the number of Z-layers and decreases with more aggressive compression. At high image quality, 10-layer datasets average ∼1 GB per slide. **c,** Representative tomograms of glandular, LSIL, HSIL, and adenocarcinoma cells compressed to yield PSNR values of 38, 40, 42, and 44 dB. At 38 dB, compression artifacts are apparent. At 40 dB and above, diagnostically important features such as nuclear membranes and nucleoli are clearly preserved. At 42 dB or higher, differences from raw images are visually negligible. Scale bars: 200 μm. **d,** Timeline of imaging operations over 10 imaging sections at a 10-layer Z-resolution. **e,** Average latency per imaging section for 10, 20, and 40 Z-layers, segmented by task (XY stage motion, image acquisition, image construction, and compression). XY motion time remains constant, while other tasks scale linearly with Z-layer count. Error bars represent standard deviation.

### Imaging performance

The whole-slide edge tomograph enables rapid, high-resolution 3D visualization of thick cytology samples, including Pap smears, sputum smears, and brush biopsy smears, as well as liquid-based cytology preparations such as ThinPrep, SurePath, and body fluid samples. The system achieves a lateral (XY) resolution of 220 nm and an axial (Z) resolution of 1 μm, generating approximately 140 gigavoxels per SurePath slide or 393 gigavoxels per ThinPrep slide. This resolution enables detailed assessment of cellular architecture, structural deformation, and abnormal morphology in any imaging plane, including oblique sections. To demonstrate its capabilities, we acquired four 3D imaging datasets at 1 μm Z-intervals, representing keratinizing and non-keratinizing squamous cell carcinoma (SCC), as well as human papillomavirus (HPV)-associated and HPV-independent adenocarcinoma. Representative tomographic slices extracted at 5 μm intervals are shown in Figures 3a-3d, showing distinct nuclear irregularity, chromatin texture, and nuclear-to-cytoplasmic (N/C) ratio shifts across SCC and adenocarcinoma subtypes – features not discernible in 2D imaging. Corresponding 3D reconstructions are shown in Figures 3e-3h, visualized along the XYZ axes to emphasize volumetric structure. Supplementary Videos 1-8 further illustrate these datasets in both 2D and 3D, providing dynamic visualizations that enhance understanding of their spatial and morphological characteristics.

**Figure 3.**
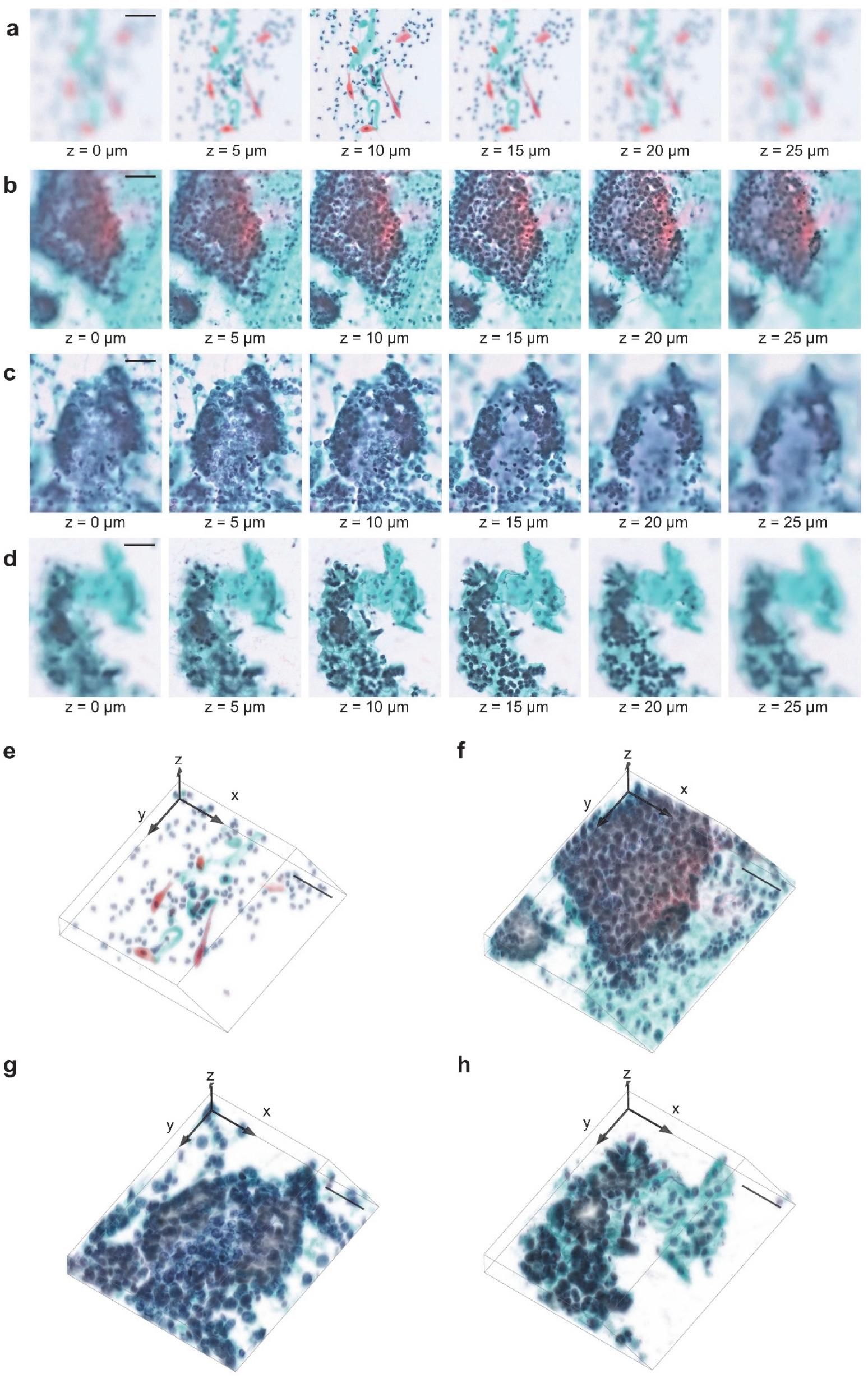
Imaging performance. **a-d,** Representative tomograms of cervical cancer cytology samples acquired at 1 μm Z-intervals and displayed at 5 μm steps. Shown are keratinizing squamous cell carcinoma (**a**), non-keratinizing squamous cell carcinoma (**b**), HPV-associated adenocarcinoma (**c**), and HPV-independent adenocarcinoma (**d**). Images highlight depth-resolved morphological features and structural deformations. **e-h,** 3D reconstructions corresponding to the samples in (**a-d**), visualized along the XYZ axes to reveal volumetric cellular architectures and spatial relationships. Scale bars: 50 μm.

### AI-based cell detection and classification performance

The availability of high-quality 3D images enables the development of a high-performance AI model for accurate cell classification. The complete workflow, from cell detection to classification, is outlined in Extended Data Figure 4. Initial 3D cell detection was performed on whole-slide tomographic images acquired at 3 μm intervals using a YOLOX-based model, trained and validated on 242,669 annotated nuclei across 348 images (Extended Data Figures 5 and 6). Detected nuclear centroids were used to extract in-focus images of individual cells, following the focus-selection procedure detailed in the Methods section. These focus-refined images were then processed by a MaxViT-based vision transformer for cell type classification. Figure 4a presents a confusion matrix comparing expert cytologist annotations with AI predictions. The model was trained and validated on a dataset of 168,569 augmented images (rotation and flipping) derived from 354 donor-based whole-slide images, with 50,222 images reserved for validation (Extended Data Figure 7). The classifier achieved high accuracy, with specificity exceeding 98% across all classes. Figure 4b shows receiver operating characteristic (ROC) curves, with AUC values greater than 0.99 for all classes except glandular cells and squamous metaplasia. Figures 4c-4k display representative tomograms and corresponding AI inference results for key gynecological cytology cell types. Figures 4c-4h include leukocytes, superficial/intermediate squamous cells, parabasal cells, squamous metaplasia, glandular cells, and miscellaneous clusters. Figures 4i-4k highlight LSIL, HSIL, and adenocarcinoma cells. These results demonstrate the model’s high sensitivity and reliability, even in the context of dense or aggregated cell populations.

**Figure 4.**
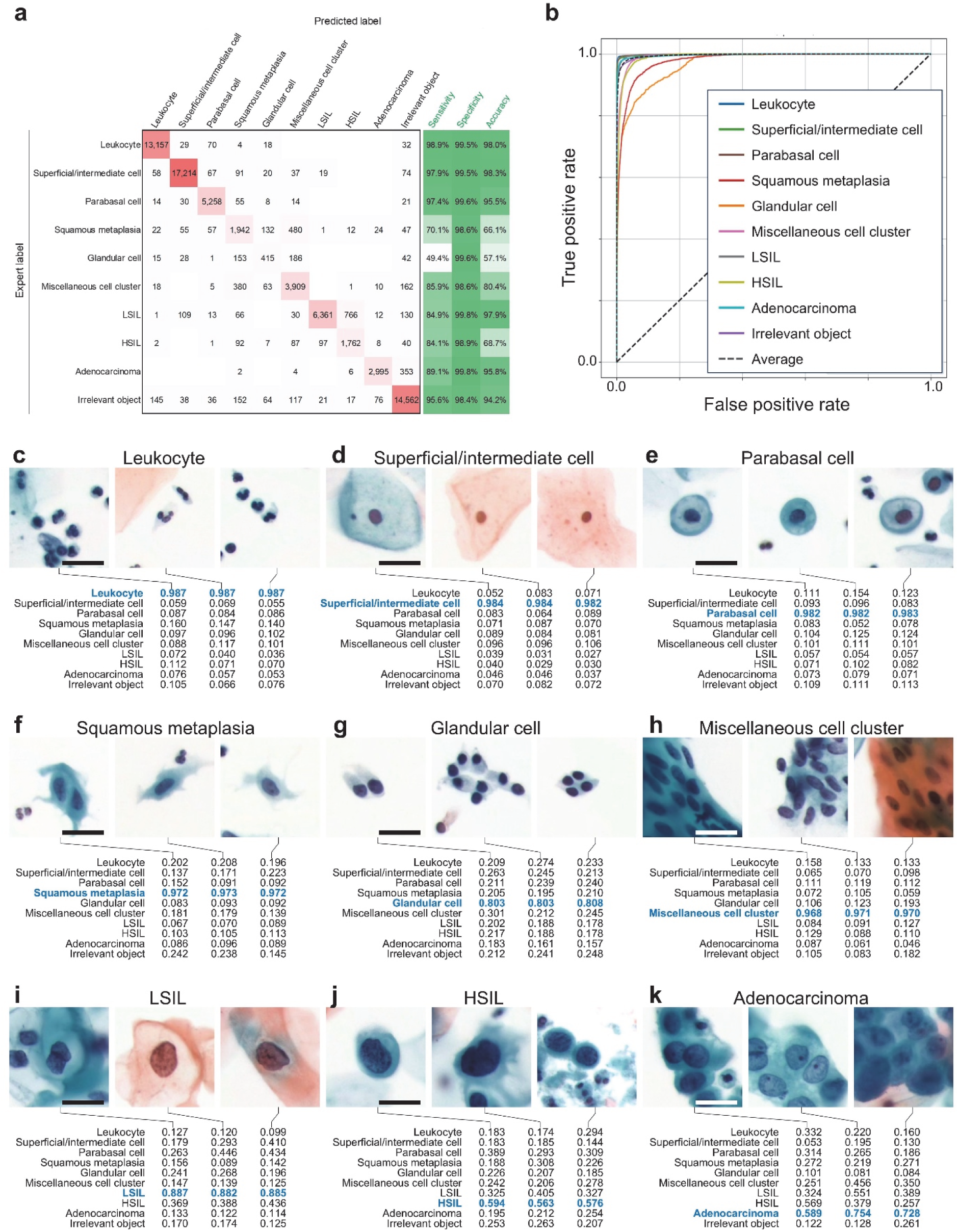
AI-based cell classification performance. **a,** Confusion matrix comparing AI inference results with expert cytologist annotations. Sensitivity, specificity, and overall accuracy metrics are provided for each cytological class. **b,** ROC curves illustrating classification performance across 10 cell types. AUC values were: 1.00 for leukocytes, superficial/intermediate squamous cells, parabasal cells, LSILs, adenocarcinoma cells, and irrelevant objects; 0.99 for HSILs and miscellaneous clusters; 0.98 for squamous metaplasia; and 0.97 for glandular cells. **c-k,** Representative AI-classified tomograms of individual cells from each category: leukocytes (**c**), superficial/intermediate squamous cells (**d**), parabasal cells (**e**), squamous metaplasia (**f**), glandular cells (**g**), miscellaneous cell clusters (**h**), LSILs (**i**), HSILs (**j**), and adenocarcinoma cells (**k**). Each panel includes the associated probability vector output by the vision transformer model, reflecting the AI’s classification confidence across all 10 categories. Scale bars: 20 μm.

### CMD-based cell population analysis

We characterized cell populations using the CMD framework and visualized them in a statistically robust and interpretable manner by incorporating AI-derived class probability values as axes in scatter plots. Figures 5a and 5b show analyses from cytological samples diagnosed as negative for intraepithelial lesion or malignancy (NILM), while Figures 5c and 5d correspond to samples diagnosed as LSIL and HSIL, respectively. Each panel includes four plots (from left to right): (1) a scatter plot for separating leukocytes and irrelevant objects, (2) a histogram of LSIL probability scores, (3) a histogram of HSIL probability scores, and (4) a uniform manifold approximation and projection (UMAP) plot. A gating strategy is applied to the scatter plots to isolate the bottom-left population, consisting primarily of epithelial cells, by excluding leukocytes and debris. The histograms are generated from this gated population, with thresholds applied to identify LSIL- and HSIL-positive cells; absolute counts and proportions are annotated. The UMAP plots visualize the spatial distribution of gated cells, with color-coded cell types enabling intuitive exploration of the cytological landscape. In Figure 5b, eight representative points (p1-p4 and q1-q4) illustrate two morphologic trajectories: p1-p4 from parabasal to superficial/intermediate squamous cells, and q1-q4 from glandular to metaplastic cells. Corresponding images are shown in Figure 5e, capturing gradual transitions in morphology and suggesting that the UMAP encodes a pseudo-histological distribution akin to in vivo tissue organization. In the UMAP plots of Figures 5c and 5d, LSIL and HSIL cells are indicated by orange and red arrows, respectively. Representative cell images from these regions, shown in Figures 5f-5h, confirm the extraction of cells exhibiting cytological atypia consistent with the corresponding diagnostic categories.

**Figure 5.**
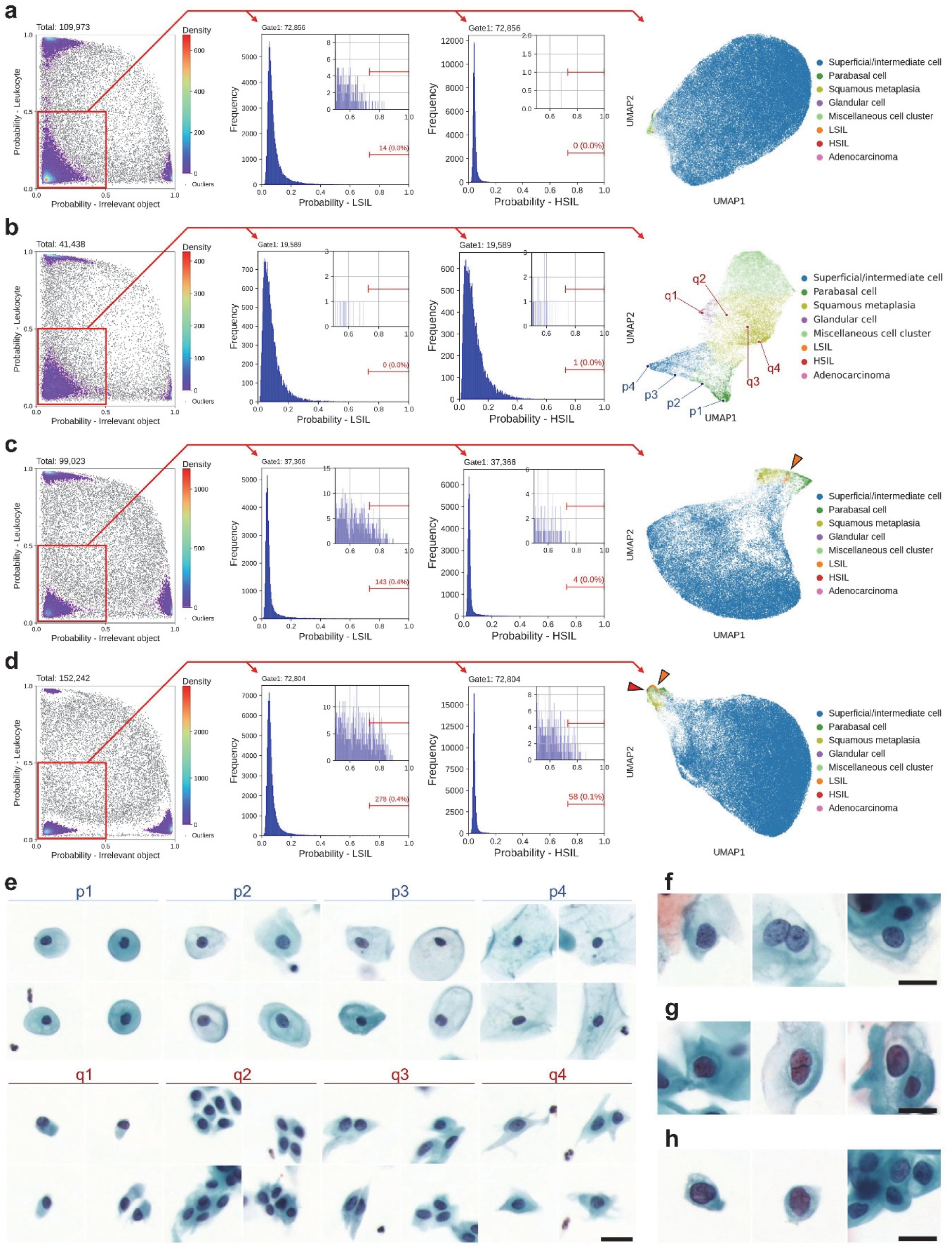
CMD-based cell population analysis. **a, b,** CMD-based analysis of cytological samples diagnosed as NILM. **c, d,** CMD-based analysis of samples diagnosed as LSIL (**c**) and HSIL (**d**). Each panel includes four plots, from left to right: scatter plot for gating out leukocytes and irrelevant objects; histogram of LSIL probability scores; histogram of HSIL probability scores; and UMAP projection visualizing the distribution of remaining epithelial cells. **e,** Representative single-cell images illustrating gradual morphological transitions observed in b: from parabasal to superficial/intermediate squamous cells (p1-p4) and from glandular to metaplastic cells (q1-q4). **f-h,** Representative images of LSIL, HSIL, and adenocarcinoma cells corresponding to the orange and red arrows in c and d, respectively. Scale bars: 20 µm.

### Clinical-grade performance

To demonstrate the clinical-grade performance of our autonomous cytology platform, comprising the whole-slide edge tomograph and the CMD-based analysis, we evaluated cervical liquid-based cytology samples from 318 donors at the Cancer Institute Hospital of JFCR. For each case, the platform quantified the number of normal, LSIL, HSIL, and adenocarcinoma cells. Figures 6a and 6b present the whole-slide cell counts by class: Figure 6a shows results for all cell types, while Figure 6b focuses on positive (abnormal) cell classes. In both figures, slides are grouped by cytology result along the horizontal axis and sorted by total cell count within each group along the vertical axis. Figure 6a reveals that total cell counts per slide vary by up to two orders of magnitude, ranging from several thousand to several hundred thousand. Slides with high cell counts typically have a higher proportion of superficial/intermediate squamous cells, indicative of robust exfoliation of superficial layers, whereas slides with lower cell counts tend to contain a greater abundance of metaplastic and parabasal cells, reflecting variations in sampling depth or patient-specific epithelial remodeling.

**Figure 6.**
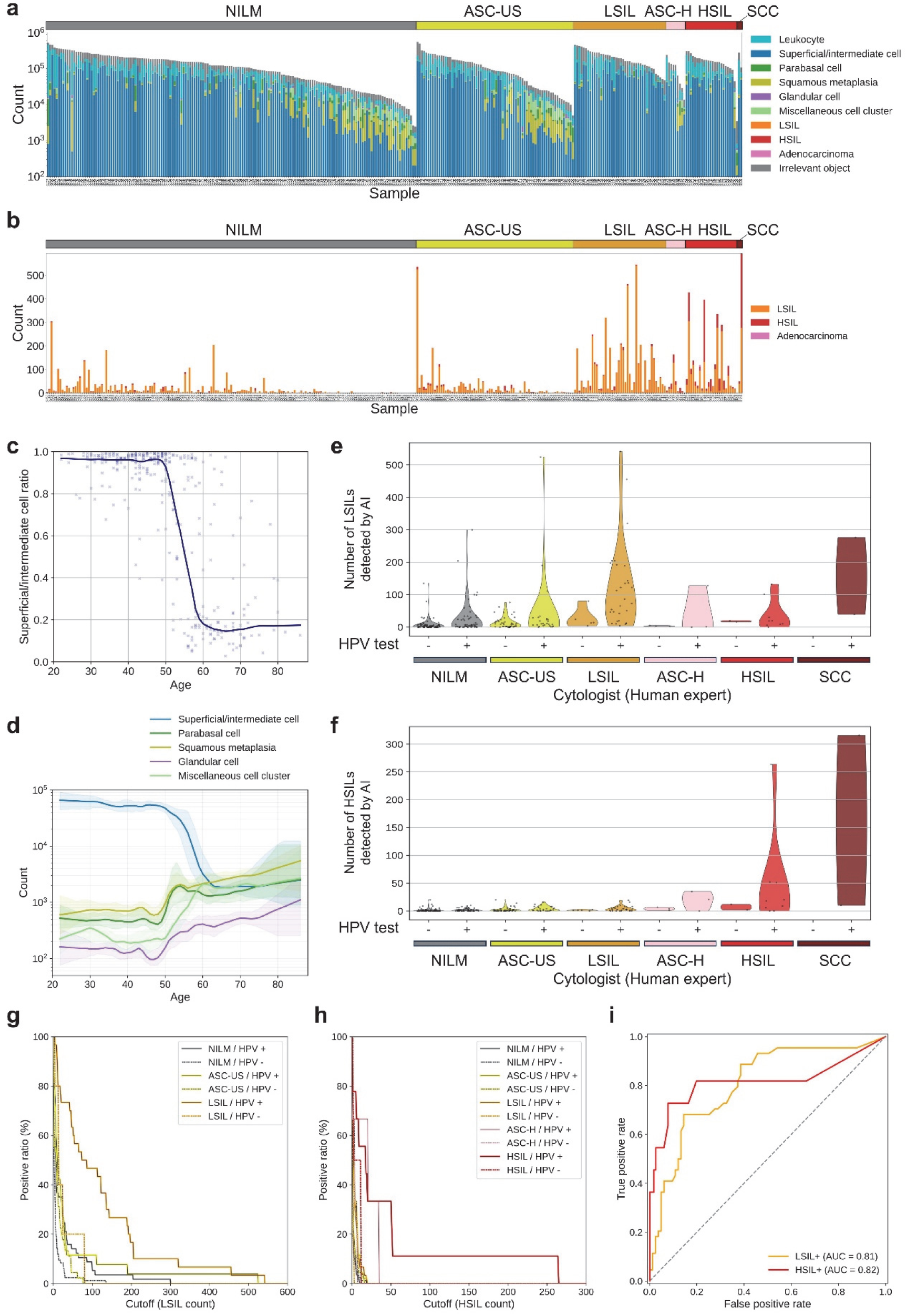
Clinical-grade performance. **a, b,** Whole-slide cell counts from 318 cervical liquid-based cytology samples, grouped by cytological diagnosis and sorted within each group by total cell count; counts across all cell types (**a**) and counts limited to abnormal (positive) cell classes (**b**). Cell counts per slide span from several thousand to several hundred thousand, with slides containing high total counts typically enriched in superficial/intermediate squamous cells. Each dot represents one slide; solid lines indicate LOWESS trends. **c,** Ratio of superficial/intermediate squamous cells as a function of donor age in NILM samples. **d,** Log-scaled absolute counts of five epithelial cell types in NILM samples by age. Shaded areas represent 95% confidence intervals based on bootstrap resampling (n = 100). **e, f,** AI-detected LSIL (**e**) and HSIL (**f**) cell counts, stratified by cytological diagnosis and HPV status. **g, h,** Proportion of slides classified as positive based on varying cell count thresholds for LSIL⁺ (**g**) and HSIL⁺ (**h**). **i,** ROC curves for detecting LSIL⁺ (LSIL, ASC-H, HSIL, SCC; orange) and HSIL⁺ (HSIL, SCC; red) cases in HPV-positive samples, using AI-derived cell counts as predictors.

Suspecting that this variation in cell composition might relate to donor age, we further investigated the ratio of superficial/intermediate squamous cells by age group (Figure 6c). This analysis was motivated by the well-established observation that epithelial turnover and differentiation are influenced by hormonal changes occurring during puberty, reproductive age, perimenopause, and postmenopause^44^, which in turn affect the overall cellular makeup of cervical cytology samples. We also examined the absolute counts of major epithelial cell types, including superficial/intermediate squamous cells, parabasal cells, squamous metaplasia, glandular cells, and miscellaneous cell clusters, across different age groups (Figure 6d). These results confirm that cell population composition changes significantly after the age of 50, with notable increases in the proportions of metaplastic and parabasal cells in older individuals. Such shifts in cellular composition may reflect age-related changes in the cervical epithelium and have important implications for the interpretation of cytological findings.

Interestingly, Figure 6b shows several slides diagnosed as NILM in which the AI model nonetheless detected large numbers of LSIL cells, suggesting possible underdiagnosis by human review or the presence of subtle HPV-related changes that were not identified by conventional cytology. Conversely, we observed some slides diagnosed as LSIL with only a few positive cells detected by the AI, indicating potential overcalls or limitations in sampling. To evaluate the validity and consistency of these cytology and AI assessments, we plotted the number of LSIL and HSIL cells detected by the AI model according to both cytology and HPV test results (Figures 6e and 6f). The AI-derived LSIL counts correlated well with HPV positivity, supporting the conclusion that the model specifically identifies HPV-associated cellular abnormalities rather than generic atypia. HSIL cell counts were elevated in cases categorized as ASC-H and above, further supporting the clinical relevance and potential diagnostic value of the AI model’s results in stratifying cervical disease severity.

To evaluate whether LSIL and HSIL positivity could be determined based on the number of respective abnormal cells, we analyzed the proportion of positive cases within each cytological category as a function of the cutoff threshold for LSIL (Figure 6g) and HSIL (Figure 6h) cell counts. In these plots, the horizontal axis represents the cutoff value used to define positivity for LSIL or HSIL cells, while the vertical axis indicates the percentage of samples classified as positive (i.e., those exceeding the specified threshold) within each cytological category. This approach allows for systematic exploration of the relationship between quantitative cell counts and cytological classification. The results showed that LSIL counts were consistently higher in HPV-positive LSIL cases compared to lower-grade samples, and HSIL counts were similarly elevated in HPV-positive HSIL and ASC-H cases relative to lower-grade samples, suggesting a clear separation of cytological severity based on these quantitative cell-count metrics.

To further assess the diagnostic utility of these cell count–based metrics, we performed ROC curve analyses on HPV-positive samples (Figure 6i). For this analysis, cases with LSIL, ASC-H, HSIL, or SCC were defined as positive for low-grade or higher-grade abnormalities (hereafter referred to as LSIL⁺), and cases with HSIL or SCC as positive for high-grade abnormalities (hereafter referred to as HSIL⁺). The sum of LSIL and HSIL cell counts was used as a score to detect LSIL⁺ cases, while the HSIL count alone was used to detect HSIL⁺ cases. The resulting AUC was 0.81 for LSIL⁺ detection and 0.82 for HSIL⁺ detection, indicating good discriminative performance for both metrics and supporting the potential of these quantitative measures to serve as reliable indicators of disease severity. These findings reinforce the value of these AI-derived cell counts as quantitative, reproducible biomarkers that complement morphology-based interpretation, particularly in borderline cytology cases such as ASC-US or LSIL without HPV testing.

## DISCUSSION

This study presents a real-time, clinical-grade, autonomous cytology platform that integrates high-speed, high-resolution whole-slide optical tomography with edge computing. By combining rapid 3D imaging, on-the-fly data compression, and population-scale morphological analysis, the system addresses long-standing challenges in cytological diagnostics – namely subjectivity, inconsistency, and limited scalability. Also, we have overcome the critical hurdle of digitizing cytology samples in conventional cytological smears and thick cell clusters in liquid-based cytology, especially given their inherent thickness^37–39^, by leveraging whole-slide edge tomography with high spatial resolution. In contrast to traditional approaches such as extended depth-of-field imaging, which often fail to resolve overlapping cells and struggle to capture structural irregularities^45,46^, our platform provides detailed 3D reconstructions that retain essential morphological information.

We have demonstrated this capability by successfully digitizing 3D structures of SCC and adenocarcinoma cells in cervical cytology smears. Notably, adenocarcinoma often presents as hyperchromatic crowded groups^47^ – densely packed structures that have traditionally posed significant challenges for digital imaging due to limited axial resolution and optical sectioning^16,48–50^. Our system addresses this by enabling high-resolution optical slicing along the Z-axis with minimal computational cost, allowing clear visualization of individual nuclei and subnuclear features within tightly clustered cells. This advancement is broadly applicable to other cytological samples from the breast, lung, thyroid, endometrium, and salivary glands^16,50^, where similarly complex 3D architectures are common. High-fidelity digitization is essential not only for remote diagnostics and educational training but also for validating AI-generated results in clinical workflows.

A central innovation of our platform is the CMD – an interpretable embedding space analogous to CD markers used in flow cytometry. The CMD enables cytologists to explore large-scale morphological landscapes using intuitive tools such as scatter plots, histograms, and UMAP projections. Through the CMD, we identified clear morphological trajectories reflecting epithelial differentiation and neoplastic transformation, offering insight into transitional states not easily captured by conventional categorical labels. Quantitative validation using silhouette scores and phenotype clustering confirmed that CMD captures biologically meaningful structure aligned with both cytological classification and HPV status.

The CMD-based quantitative cytology framework offers significant potential for advancing diagnostic precision and screening efficiency. As shown in Figure 6i, the system successfully distinguished LSIL and HSIL cells based on their abundance in HPV-positive samples, suggesting that CMD-based cytology could serve not only as a supplement to primary screening but also as a triage tool for stratifying HPV-positive individuals. Importantly, previous studies have highlighted that human interpretation in cytology can be biased by knowledge of HPV status, potentially affecting diagnostic outcomes^51–53^. In contrast, our AI-driven quantitative approach delivers consistent and objective assessments, thereby reducing such bias and enhancing diagnostic reliability across diverse clinical settings.

Beyond classification, the CMD enables high-dimensional phenotypic analysis. As illustrated in Figures 5c and 5d, UMAP trajectories capture gradual morphological transitions, offering insight into spatial and temporal cell-state dynamics, including potential disease progression. Figures 6c and 6d further reveal age-associated shifts in epithelial composition, underscoring the CMD’s utility for population-level cytological profiling. Notably, Figure 6e shows that in NILM samples, the AI model identified a higher number of abnormal cells in HPV-positive cases than in HPV-negative ones – cells that may have been overlooked by human screeners. As shown in Figures 6a and 6b, these cases often exhibited high overall cell counts and a predominance of superficial/intermediate squamous cells – conditions that can overwhelm manual reviewers and hinder detection of rare abnormalities. The AI model’s ability to detect such subtle patterns highlights a key advantage of CMD-based, large-scale single-cell analysis. Collectively, these findings demonstrate the power of CMD-based AI cytology not only as a diagnostic aid but also as a discovery tool for uncovering latent patterns in complex cellular populations.

While further development and validation are needed, our system presents significant potential to enable a new generation of applications in cytological diagnostics and research. First, the integration of 3D digitization with AI-based analysis could help mitigate the global shortage of cytotechnologists^54–56^ by streamlining diagnostic workflows and expanding access to rapid on-site procedures such as ROSE^57^, which are currently restricted to specialized centers. Second, the system can extend cytological diagnostics to rural and underserved regions where trained cytologists are unavailable, thereby improving access to timely and reliable care. Third, by transforming cellular morphology into quantitative and reproducible metrics, this approach can facilitate knowledge transfer, reduce dependence on subjective expertise, and accelerate the discovery of novel cell types or diagnostic markers. Finally, coupling this system with technologies such as image-activated cell sorting^58,59^ could enable real-time isolation of rare or abnormal cells based on subtle morphological features. This would pave the way for downstream molecular analyses, linking cytological imaging directly to genomics and proteomics. Together, these advances point toward a more accessible, autonomous, and data-driven future for cytology.

## METHODS

### Whole-slide edge tomograph

As shown in Extended Data Figure 1, the whole-slide edge tomograph comprises multiple hardware modules optimized for high-speed 3D imaging and edge-side data processing. The illumination system utilizes a high-power LED (XQ-E, Cree) as the light source, paired with a motorized iris (Nihon Seimitsu Sokki Co., Ltd., Japan) to control the numerical aperture. This illumination passes through the cytology sample and is projected onto a camera board equipped with a CMOS image sensor (IMX531, Sony) and imaging optics. The camera board (e-con Systems, India) is mounted on a Z stage (Chuo Precision Industrial Co., Ltd., Japan), which executes precise axial scanning under the control of a real-time controller. The XY stage translates the slide in the lateral plane for complete coverage during image acquisition.

These mechanical components are tightly integrated with the edge computer, which includes several modules: an image sensor FPGA (CertusPro-NX, Lattice Semiconductor), a real-time controller equipped with an additional FPGA (Artrix7, Advanced Micro Devices, Inc.), an XY stage controller based on a microcontroller (STM32, STMicroelectronics), an illumination controller based on a microcontroller (RL78, Renesas Electronics), and a system-on-module (SOM) unit (Jetson Xavier NX, NVIDIA). The SOM features a multi-core CPU, a GPU, a hardware encoder, and main memory used as an image buffer. An application running on the SOM manages internal communications over USB serial and SPI protocols to coordinate the XY stage, Z stage, and illumination modules. Captured images from the CMOS sensor are first transmitted to the FPGA, where real-time high-speed signal conditioning and protocol conversion are performed.

To support high spatial and temporal resolution, the system utilizes a dual 4-lane MIPI interface, which effectively doubles the data throughput compared to a single MIPI-CSI configuration. This allows continuous transmission of 4.5k × 4.5k resolution images at up to 50 frames per second from the FPGA to the SOM, facilitating reliable real-time handling of large volumetric datasets. Upon receipt by the SOM, image data undergo a three-step processing pipeline: (1) 3D image acquisition, (2) 3D reconstruction via axial alignment using both the GPU and CPU, and (3) real-time compression using the on-board encoder. For compression, the system leverages NVIDIA’s NVENC library to encode 3D image stacks into the HEVC format with hardware acceleration. This process ensures substantial data reduction while maintaining the critical structural features needed for downstream visualization and analysis. The resulting compressed image data are stored locally on a solid-state drive (SSD) integrated within the edge computer.

From there, compressed image data are transmitted to a backend server, where they are stitched into full-slide 3D volumes and stored on a network-attached storage (NAS) system. These reconstructed volumes are subsequently used for both interactive visualization and AI-based computational analysis. The backend server hosts a DZI viewer, which enables smooth and responsive visualization by dynamically decompressing and transmitting only the requested tile regions based on user inputs such as zooming, panning, and focus adjustments. These operations are accelerated by a GPU (RTX 4000 Ada, NVIDIA), which handles stitching, image rendering, and hardware-accelerated decoding. In parallel, an AI analysis server retrieves the compressed data from the NAS, decodes it using hardware acceleration, and performs diagnostic or morphological inference using a high-performance GPU (RTX 6000 Ada, NVIDIA). The resulting predictions and associated metadata are stored back on the NAS for subsequent review or downstream integration.

### Sectional 3D image construction and compression

The imaging workflow involves tightly coordinated real-time interactions among multiple software and hardware components operating in parallel. The real-time controller adjusts the Z stage to sequentially position the slide at specified focal depths, enabling the acquisition of sectional 2D images across various Z-planes. Concurrently, each captured image is transmitted to the image signal processing unit of the FPGA, which forwards the data to the GPU buffer on the edge computer. Upon completion of image acquisition at a given region, the XY stage promptly moves the slide to the next imaging section, while image processing and compression begin immediately – achieving a pipelined, non-blocking execution flow. A dedicated 3D image construction module processes the acquired Z-stack by enhancing color uniformity and dynamic range and selecting optimal focal planes to ensure that all cells appear sharply focused. In parallel, a 3D image compression module utilizes the hardware encoder integrated in the SOM to compress the processed image stack into an HEVC-format video file. These modules operate simultaneously, enabling high-throughput scanning without computational bottlenecks.

To evaluate the timing characteristics of individual imaging tasks, time logs were recorded under three Z-layer configurations: 10, 20, and 40 layers. Representative task sequences for the first 8 imaging sections at the beginning of a whole-slide scan are shown in Figure 2d (10 layers), Extended Data Figure 2a (20 layers), and Extended Data Figure 2b (40 layers). These logs delineate task execution for XY stage motion, image acquisition, 3D construction, and compression. The first imaging section includes an initialization step and thus takes slightly longer than subsequent areas.

To quantify system performance across the full scan, we calculated the average and standard deviation of each task’s latency per imaging section for each Z-layer setting. As summarized in Figure 2e, XY stage motion time remained constant regardless of Z-stack depth, while image acquisition, construction, and compression durations increased linearly with the number of Z-layers. The larger error bars associated with XY stage motion reflect variations in travel distance between imaging sections. These results confirm the system’s predictable and efficient scaling behavior across varying imaging depths.

### Sectional 3D image compression

Sectional 3D image compression was implemented using the HEVC codec, with optimized parameters defining three selectable modes: high, medium, and low, corresponding to target bitrates of 40.36, 24.21, and 8.07 Mbps, respectively. To evaluate compression performance, we assessed both image quality and resulting file size. Image quality was quantified by calculating the PSNR between original and compressed YUV images. For each compression setting, three cytology slides were scanned, with 10 imaging sections per slide and 10 Z-layers per area, yielding 300 frames per setting. PSNR values were computed for every frame and visualized as histograms in Figure 2a.

To analyze frame-wise variation in compression artifacts, we further plotted PSNR values across Z-layers for each slide and imaging section combination (30 lines per setting). These Z-layer-wise profiles, shown in Extended Data Figure 2c, revealed that under low-quality settings, PSNR values exhibited an alternating pattern between even and odd layers – an effect that diminished at higher bitrates. In many imaging sections under the low setting, PSNR fluctuated around 41-42 dB, whereas medium and high settings showed more stable PSNR levels centered around 42 dB and 43 dB, respectively. We also noted that some imaging sections exhibited higher-than-average PSNR values across all conditions. Upon inspecting the corresponding regions in the original slides, we found that these locations coincided with red ink markings manually applied to the coverslip. Since these markings present simpler and more uniform visual features than the surrounding cellular structures, they likely resulted in less distortion during compression, yielding higher PSNR.

To investigate the correlation between PSNR and visual acceptability for cytological interpretation, we generated compressed image sets with finely adjusted compression parameters that produced PSNR values ranging from 38 to 46 (Figure 2b). Visual inspection showed that image degradation was negligible when PSNR exceeded 40 dB, and virtually imperceptible above 42 dB. Since the predefined High and Medium compression modes consistently yielded PSNR values around 42 and above 40, respectively, we concluded that these settings maintain sufficient fidelity for reliable cytological assessment.

We next examined how compression quality settings influence processing time (Extended Data Figure 2d). Under the 10-layer condition, we compared time distributions across High, Medium, and Low compression modes for each task. As expected, XY stage motion, image acquisition, and image construction times remained unaffected by compression settings. Image compression time increased slightly with higher quality settings, but the increase was modest and did not significantly impact overall system throughput. These results confirm that high-quality image compression can be achieved without compromising imaging efficiency.

Finally, we evaluated file size and imaging time using three additional cytology slides, independently from those used for PSNR analysis. Whole-slide 3D images were acquired under three compression quality levels (high, medium, low) and Z-layer counts (10, 20, 40). Resulting data sizes are shown in Figure 2c. As expected, file size increased proportionally with the number of Z-layers and decreased with stronger compression. Imaging time under these same conditions was measured separately and is presented in Extended Data Figure 2e. These values represent the duration of the pure image acquisition process, excluding slide loading or system preparation time. Imaging time scaled linearly with the number of Z-layers and was not substantially affected by compression settings.

### Sectional 3D image decompression and viewing

Extended Data Figure 3a illustrates the architecture of the DZI viewer system, which enables interactive, web-based visualization of whole-slide 3D cytology images. The system is composed of three primary layers: the frontend, backend, and data layer. Following image acquisition, sectional 3D images are transmitted from the imaging system to the backend server over a network. The backend software then performs stitching of individual imaging sections using positional metadata, generating the alignment information required to reconstruct the full 3D whole-slide image. Once stitching is completed, the image data are made available for viewing through the DZI interface.

In the frontend, users can browse a list of available slides and interact with the whole-slide image using familiar operations such as zooming, panning, rotation, and navigation across Z-layers. A preview image assists with rapid slide identification, and annotation tools allow users to flag or mark suspicious cells. Crucially, the ability to scroll through focal planes enables inspection of diagnostically relevant features that may be missed in conventional 2D views.

On the backend, the server responds to frontend requests via two application programming interfaces (APIs): a slide API for metadata and an image API for tile access. When a specific image tile is requested, the backend retrieves the corresponding compressed frame from the data layer and decompresses it using NVDEC, a hardware video decoder integrated into the NVIDIA GPU. The Decord library interfaces with NVDEC, enabling efficient, hardware-accelerated HEVC frame decoding and supporting random access for rapid tile retrieval. The system is capable of handling more than ten concurrent image tile requests in parallel, ensuring responsive, low-latency performance even under high user load.

The performance of the sectional 3D image decompressor was evaluated to assess the backend server’s ability to respond to image requests from the viewer in real time. A series of tests measured the response time as a function of request frequency and image tile size, quantifying the system’s capability to retrieve and render tomograms efficiently under varying conditions. The evaluation involved benchmarking the time required to retrieve and display tomographic frames. The results, presented in Extended Data Figure 3b, show that the system maintained an average response time of under 100 msec per image tile request, even when on-the-fly HEVC decompression was required. Integration of NVDEC interfaces significantly reduced decompression latency, while the Decord library enabled fast, hardware-accelerated frame extraction. These findings confirm the system’s efficiency and scalability, demonstrating its suitability for high-throughput, real-time applications in medical image analysis, where low latency and responsiveness are critical for clinical utility.

### Detection of cell nuclei

Cell nuclei were detected using a YOLOX object detection model trained on a cytology-specific dataset derived from images acquired by the whole-slide edge tomograph (Extended Data Figure 2a). A total of 348 images were used, with 278 allocated for training and 70 for validation. Initial nucleus annotations were generated using a semi-automated pipeline based on traditional image processing methods, and then manually reviewed and corrected using the computer vision annotation tool (CVAT). This process yielded 242,669 annotated nuclei in total; 199,552 for training and 43,117 for validation (Extended Data Figure 5).

To reduce computational overhead, both training and inference were conducted on downsampled images resized from the original 4.5k × 4.5k resolution to 1k × 1k pixels. Model training was performed on an NVIDIA RTX A6000 Ada GPU. Inference performance was evaluated using ROC curve analysis across intersection-over-union (IoU) thresholds of 0.8, 0.6, 0.4, and 0.0, yielding AUC values of 0.79, 0.77, 0.70, and 0.63, respectively (Extended Data Figure 6a). To maximize sensitivity and minimize missed detections during this critical initial stage, we selected an IoU threshold of 0.0 and a detection probability cutoff of 0.005.

To assess detection validity, we compared automated nucleus counts to manual counts across the validation dataset. As shown in Extended Data Figure 6b, the two methods exhibited strong agreement, with a regression line of y = 1.0098x and R² = 0.9487, indicating high correlation. Outlier cases, where the model overestimated counts, were further examined (Extended Data Figures 6c and 6d). These discrepancies were largely attributed to non-specific detections near cell boundaries or in background regions. However, such false positives were deemed acceptable, as downstream cell classification processes are designed to filter out irrelevant detections. In contrast, false negatives at this stage would result in the exclusion of cells from subsequent analysis, which would be more detrimental to overall performance.

### Extraction of in-focus single-cell images centered on nuclei

To extract morphologically informative single-cell images, we employed a multi-step pipeline involving nucleus detection, Z-layer grouping, focus evaluation, and image cropping (Extended Data Figure 2b). Initial nucleus detection was performed using the YOLOX model on downsampled versions (1k × 1k pixels) of the original 3D whole-slide images. To balance axial resolution and inference speed, 2D optical sections were subsampled at 3 μm intervals from the Z-stack. This approach often resulted in multiple redundant detections of the same nucleus across neighboring slices, due to partial visibility in adjacent planes.

To resolve this redundancy, we implemented a grouping algorithm that clustered spatially proximate and axially aligned detections across Z-layers, treating them as a single nucleus instance. For each grouped nucleus, we identified its Z-range and retrieved full-resolution (4.5k × 4.5k pixels) image patches centered at the nucleus coordinates, but only within the identified Z-range. This selective retrieval ensured that focus evaluation was conducted on the relevant high-resolution slices, minimizing unnecessary computation.

A focus evaluation metric was then applied to the extracted Z-stack to identify the slice with the best optical focus. The slice with the highest focus score based on criteria such as local contrast or sharpness was selected. From this slice, a 224 × 224 pixel patch centered on the nucleus was cropped, capturing both the nucleus and its surrounding cytoplasmic context. These high-quality, nucleus-centered single-cell images served as inputs for downstream classification models.

### Classification of single-cell images using MaxViT

Single-cell images extracted from the focused optical section of each nucleus-centered region were classified using a vision transformer model based on the MaxViT-base architecture. The model was trained to differentiate among ten cytological categories: leukocytes, superficial/intermediate squamous cells, parabasal cells, squamous metaplasia cells, glandular cells, miscellaneous cell clusters, LSIL, HSIL, adenocarcinoma cells, and irrelevant objects (e.g., debris, non-cellular material, and defocused images). The training and validation datasets were constructed from expert-annotated cell images derived from 354 donor-derived whole-slide images. The number of annotated cells used for training and validation in each class was as follows: 18,219 / 5,281 leukocytes; 23,158 / 7,557 superficial/intermediate squamous cells; 4,296 / 1,243 parabasal cells; 2,056 / 487 squamous metaplastic cells; 836 / 105 glandular cells; 5,387 / 994 miscellaneous cell clusters; 1,846 / 936 LSIL cells; 1,433 / 262 HSIL cells; 912 / 420 adenocarcinoma cells; 14,752 / 5,115 irrelevant objects. All annotations were performed by professional cytologists. To improve generalization performance, the dataset was augmented using random rotations and horizontal/vertical flipping. This resulted in a total of 168,569 training images and 50,222 validation images. Representative examples of the training images are shown in Extended Data Figure 7. Model training was performed using an NVIDIA RTX A6000 Ada GPU.

### CMD-based cell population analysis

For each classified cell image, the vision transformer model outputs a 10-dimensional (10D) vector of class probabilities, referred to as the CMD values. These CMD vectors are computed by applying a sigmoid function to the final layer’s logits of the MaxViT model, converting them into values between 0 and 1 for each of the ten cytological classes. Each value represents the model’s confidence level for assigning a cell to a given class. Representative CMD outputs are visualized alongside corresponding cell images in Figures 4c-4k.

To analyze cell populations at the whole-slide level, we performed a series of visualization and gating-based analyses using the CMD vectors of all detected cells (Figures 5a-5d). The analysis began with a 2D scatter plot using the CMD probabilities for the “irrelevant object” and “leukocyte” classes. This enabled negative gating to exclude non-cellular, defocused, or leukocyte-dominated regions and to isolate epithelial-lineage cells. Within this gated population, we generated histograms of CMD values for the LSIL, HSIL, and adenocarcinoma classes to evaluate lesion-associated probability distributions. For each histogram, class-specific thresholds were applied to identify cells with high classification confidence, allowing sensitivity tuning for detecting abnormal populations.

Before UMAP visualization, each cell was assigned a class label using a hierarchical rule-based decision process. Cells falling beyond the irrelevant-object threshold in the scatter plot were first labeled as “irrelevant.” Of the remaining cells, those exceeding the leukocyte threshold were labeled as “leukocytes.” Among the rest, any cell that crossed a predefined gate in the LSIL, HSIL, or adenocarcinoma histograms was labeled accordingly. Cells that did not meet any lesion threshold were assigned to the class with the highest CMD value among the six remaining categories: superficial/intermediate squamous cells, squamous metaplasia, parabasal cells, glandular cells, and miscellaneous clusters.

These class assignments were then used to color-code cells in UMAP plots, with each point representing a single cell. Alpha transparency was modulated to reflect prediction confidence, enhancing visual interpretability. The resulting UMAP embeddings and class labels served as the basis for all subsequent whole-slide cytological analyses.

### Human subjects

We analyzed 770 cervical cytology samples, including 766 liquid-based cytology samples and 4 conventional smear samples, collected from 770 patients who underwent cervical cytology testing at the Cancer Institute Hospital of the Japanese Foundation for Cancer Research (JFCR) between 2011 and 2019. The study was approved by the Institutional Review Board of JFCR (IRB No. 2019-GA-1190) and conducted in accordance with the principles of the Declaration of Helsinki. Informed consent was obtained from all participants through an opt-out process.

### Sample preparation and evaluation

Cervical cells were collected using a Bloom-type brush (J fit-Brush, Muto Pure Chemicals Co. Ltd.). Liquid-based cytology samples were prepared using one of the following methods: ThinPrep (Hologic), or SurePath (Becton Dickinson and Co.), in accordance with the manufacturer’s instructions. The samples were subsequently stained using the Papanicolaou method and evaluated by cytotechnologists based on the Bethesda System for Reporting Cervical Cytology^60^. In parallel with cytological examination, HPV testing was conducted using the Hybrid Capture 2 (HC2) assay (Qiagen). The same ThinPrep or SurePath sample was used for this test. The HC2 test was carried out strictly according to the manufacturer’s protocol.

### Procedure for clinical-grade performance evaluation

For the clinical-grade performance analysis, the cervical liquid-based cytology samples from 318 donors were analyzed. Expert cytological diagnoses and HPV test results had been obtained in advance, as described elsewhere. Whole-slide image acquisition and CMD-based classification were conducted using the methods previously described, and the same classification gates defined in Figures 5a-5d were applied in this analysis. AI-based classification results were aggregated on a per-slide basis to compute the number of detected objects in each class. Figure 6a presents the total cell counts across the ten classes, including epithelial cell types and irrelevant objects, while Figure 6b focuses specifically on abnormal cell classes. In both figures, samples are grouped by cytological diagnosis and sorted within each group according to total cell count.

To investigate age-related variation in epithelial composition, the ratio of superficial/intermediate squamous cells was calculated for samples diagnosed as NILM and plotted against donor age (Figure 6c). Additionally, absolute counts of five cytologically normal epithelial components (i.e., superficial/intermediate squamous cells, parabasal cells, squamous metaplastic cells, glandular cells, and miscellaneous clusters) were calculated and plotted as a function of donor age on a logarithmic scale, with LOWESS smoothing applied. The shaded regions in Figure 6d represent 95% confidence intervals estimated through bootstrap resampling (n = 100).

The number of LSIL and HSIL cells detected by AI for each slide was visualized using violin plots (Figures 6e and 6f), with samples grouped by cytological diagnosis and HPV test results. To assess the relationship between abnormal cell counts and diagnostic categories, we calculated the proportion of slides that exceeded various cutoff thresholds for LSIL⁺ and HSIL⁺ classification within each cytological group (Figures 6g and 6h). LSIL⁺ was defined as cases diagnosed as LSIL, ASC-H, HSIL, or SCC, while HSIL⁺ was defined as HSIL or SCC. ROC analysis was performed on HPV-positive samples using the same diagnostic criteria. The total count of LSIL and HSIL cells was used for LSIL⁺ detection, and the HSIL count alone for HSIL⁺ detection. The resulting area under the curve (AUC) values are shown in Figure 6i.

## Data availability

The cytology datasets analyzed in this study are securely maintained by CYBO to safeguard patient privacy and proprietary imaging data. Due to ethical and regulatory constraints, these datasets are not publicly available. Academic investigators with no relevant conflicts of interest may request access to selected cytological features for non-commercial, research-only purposes, subject to review and approval by both CYBO and the Cancer Institute Hospital of JFCR. All data recipients must agree not to redistribute the data. Requests should be directed to N.N. at. CYBO reserves the right to decline requests at its discretion and will respond within one month of receipt.

## Code availability

Portions of the source code developed for this study, including modules for edge-device image processing, AI model training, and backend infrastructure, contain proprietary components and are not publicly released to protect intellectual property. However, source code for downstream data analysis and figure generation is publicly available and can be accessed at: https://github.com/KK-CYBO/cyboscan-manuscript.

## Acknowledgements

This work was supported by funding from the Tokyo Metropolitan Small and Medium Enterprise Support Center, the Japan Agency for Medical Research and Development (AMED; Grant Number: JP20he102202), the Ichimura Foundation for New Technology, and Koto City. We also acknowledge support from the Advanced Medical Device Acceleration Project of Tokyo Prefecture, as well as from NVIDIA Inception, Microsoft for Startups, AWS Startups, Google for Startups, and Serendipity Lab. We are grateful to Incubate Fund and JAFCO for their financial support. We further thank Akira Mishima, Takeichiro Sekiya, Minoru Oikawa, Miki Kanematsu, Noriyuki Furuta, Junzo Fujiyama, and Shigeko Yamazaki for their valuable assistance.

## Author information

### Contributions

N.N. and T.S. conceptualized the whole-slide edge tomograph. N.N. and Y.M. were responsible for designing and constructing the optical and mechanical hardware, while T.S. developed and built the electrical system. T.S., M.H., and J.Z. designed, implemented, and evaluated the software for the edge computer, including image compression. N.N., T.S., A.J., and P.K. developed, implemented, and assessed the software for image decompression, stitching, and real-time tomogram review. N.N., T.S., and R.U.I. performed data preparation and AI model training and were responsible for designing, implementing, and evaluating the software for nucleus detection, focusing, classification, and population analysis. Y.S., T.I., K.I., H.A., and T.C. prepared and reviewed the cytology samples. Y.S., T.I., K.I., H.A., S.I., and N.H. annotated the cytology data. N.N., Y.S., T.S., and T.C. planned and conducted the clinical study with gynecological samples. N.N., Y.S., T.S., K.G., and T.C. supervised the overall work. N.N. and K.G. supervised and validated the scientific aspects of the whole-slide edge tomograph. N.N., Y.S., T.S., R.Y.O., K.G., and T.C. contributed to writing the manuscript, with support from T.D. and Y.L. in preparing figures and supplementary materials. All authors were involved in editing the manuscript.

### Inclusion and ethics

This study followed ethical guidelines with informed consent obtained for all samples and protocols approved by institutional ethics committees. Data were analyzed with awareness of potential biases. We are committed to promoting equity and inclusion in research while advancing scientific understanding.

### Ethics declarations

N. N., T. S., and K. G. are shareholders of CYBO. N. N., T. S., and K. G. are inventors of patents covering the data analysis and display method. N. N. and T. S. are inventors of patents and patent applications covering the 3D image compression and processing techniques. The other authors have no competing interests.

---

**Extended Data Figure 1.**
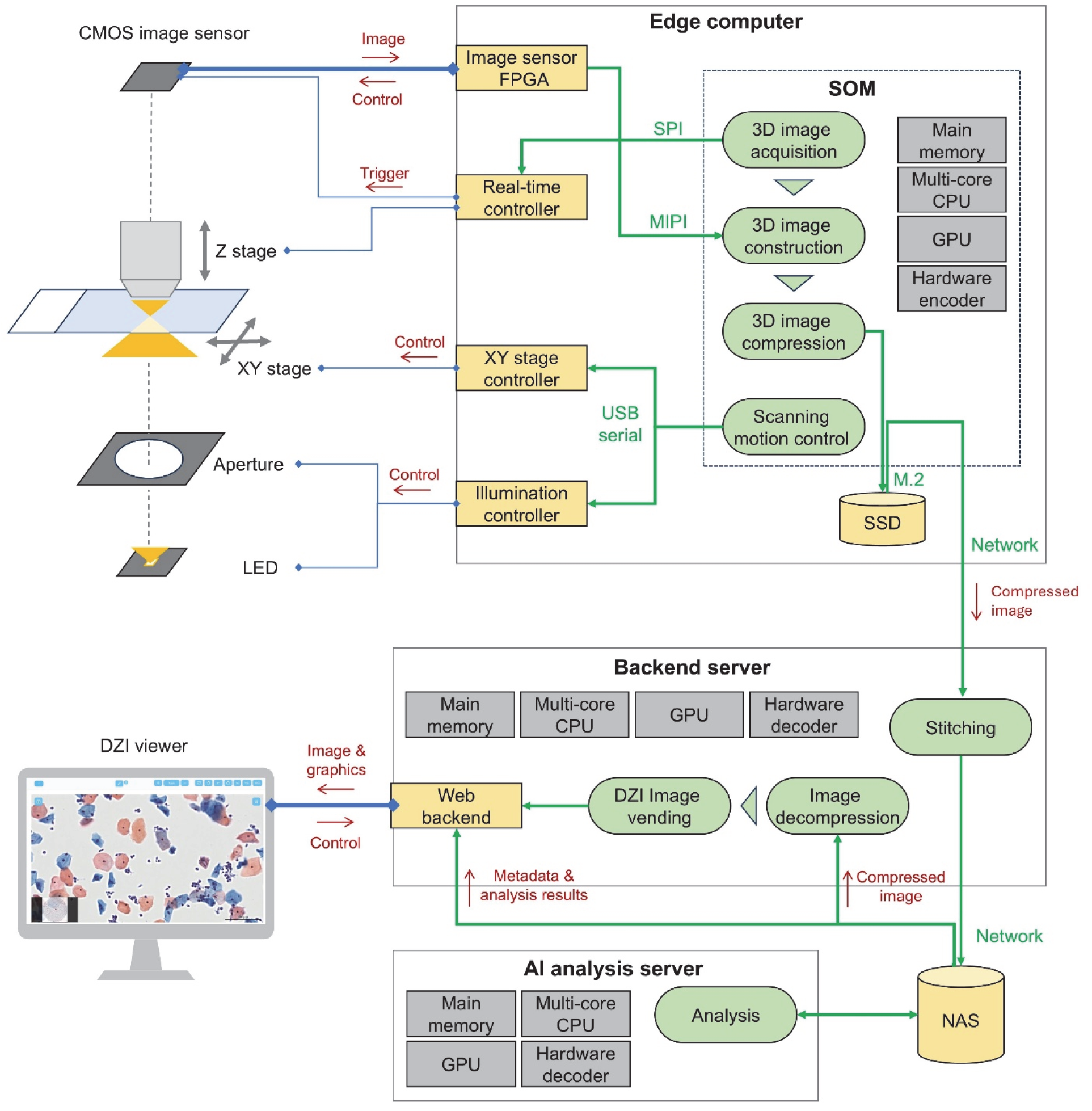
Architecture of the whole-slide edge tomograph. The system consists of an integrated hardware-software platform that enables high-speed 3D image acquisition, reconstruction, and compression directly on an SOM. The SOM coordinates with the image sensor, XY stage, and illumination controller to perform synchronized scanning and 3D imaging, with real-time processing and compression executed via its onboard GPU and hardware encoder. Scanning motion is interleaved with image acquisition to ensure full-slide coverage. Compressed image data are transmitted over the network to a backend server, where stitching is performed and results are archived in NAS. An AI analysis server retrieves the compressed data, performs decoding and inference, and stores outputs alongside image metadata. The backend also supports a DZI viewer that dynamically decompresses and streams image tiles in response to user actions such as zooming, panning, and Z-plane navigation, enabling low-latency, interactive visualization without requiring full-volume decompression.

**Extended Data Figure 2.**
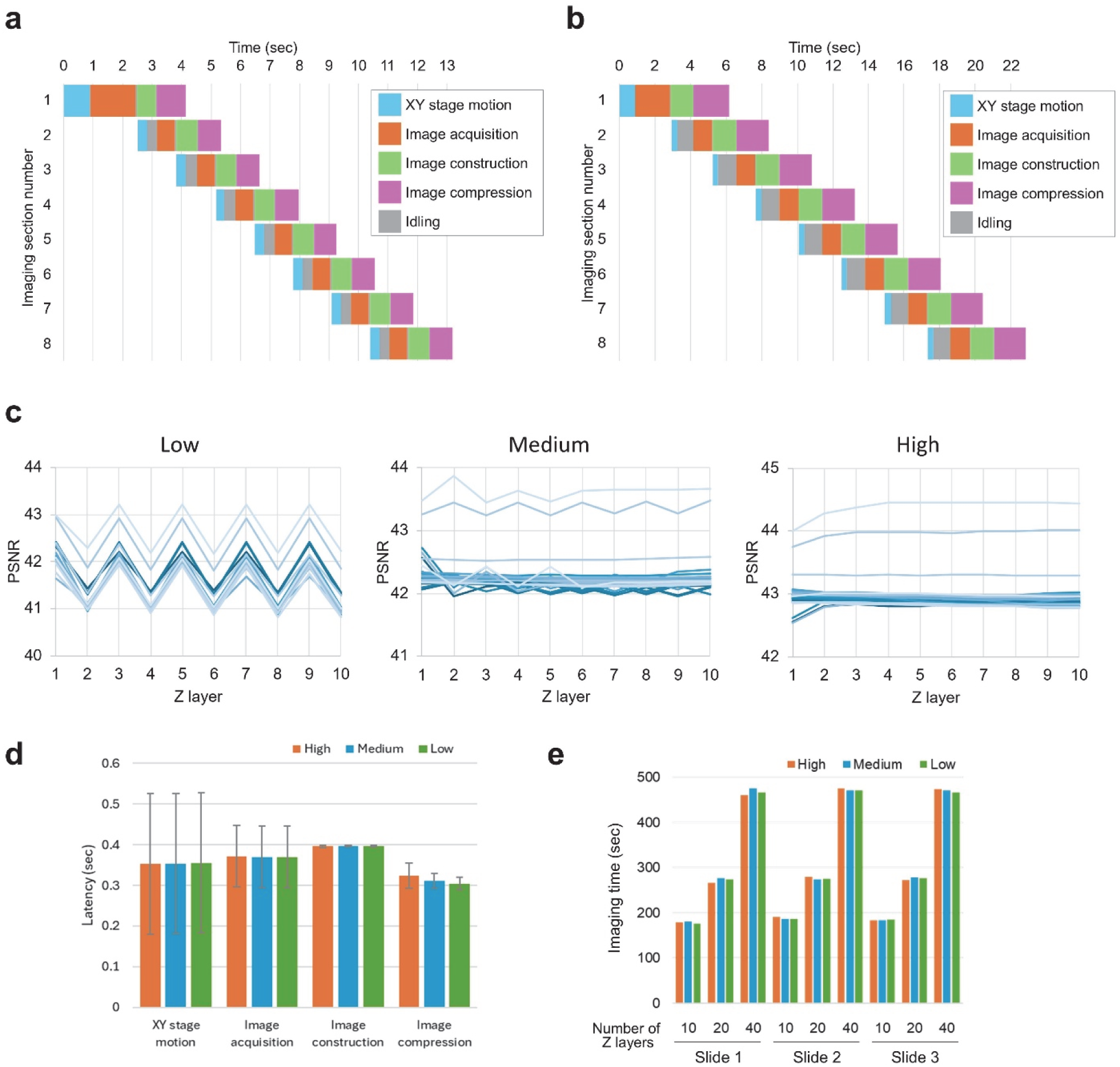
Imaging speed of the whole-slide edge tomograph. **a, b,** Time logs of the imaging process from the first to the tenth imaging section under 20-layer (**a**) and 40-layer (**b**) Z-stack conditions. Each task is color-coded: XY stage motion, image acquisition, image construction, image compression, and idle time. **c**, Line plots of PSNR values across Z-layers for each of the 30 combinations of slide and imaging section (3 slides × 10 imaging sections), based on the same 300 images analyzed in Figure 2a. Each line represents one imaging section. The three panels (left to right) correspond to low, medium, and high compression quality. Under low compression conditions, a distinct alternating pattern in PSNR values between even and odd Z-layers is observed, indicating non-uniform compression effects across depth. **d,** Average latency per imaging section for each imaging task (i.e., XY stage motion, image acquisition, image construction, and image compression) measured under high, medium, and low HEVC compression settings with 10 Z-layers. Error bars indicate the standard deviation across imaging sections. **e**, Durations for whole-slide 3D image acquisitions across varying numbers of Z-layers (10, 20, 40) and compression settings (high, medium, low). Reported values represent net acquisition time, excluding preparatory steps such as slide loading and system initialization.

**Extended Data Figure 3.**
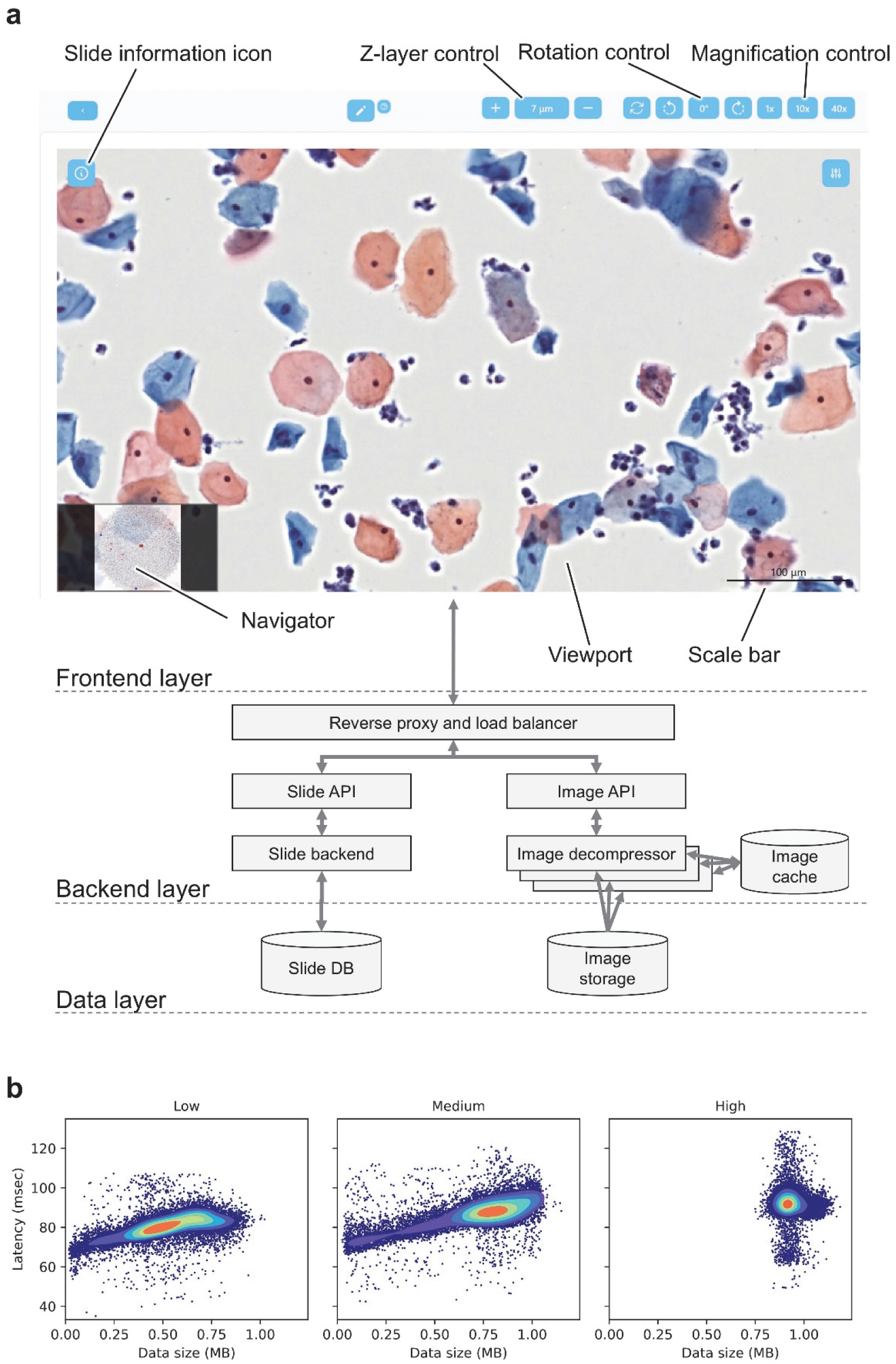
Architecture and performance of the DZI viewer. **a,** User interface and system architecture of the DZI viewer. The frontend displays whole-slide tomographic images with associated sample metadata and supports interactive operations such as zooming, XY panning, and Z-layer navigation. The backend processes client-side requests, retrieves image tiles and metadata from compressed storage, performs hardware-accelerated or GPU-based decompression, and serves the data via the image API. The system architecture is modularly organized into frontend, backend, and data layers. **b,** Scatter and density contour plots illustrating frontend request latency for DZI image tiles at the highest resolution level. The dataset includes two slides acquired under three image quality settings (low, medium, high) and three Z-layer configurations (10, 20, 40). Under low and medium compression, tile sizes vary substantially and exhibit a positive correlation with latency. In contrast, high-quality settings yield more uniform and generally larger tile sizes. Despite the computational overhead of HEVC decompression, the majority of requests are fulfilled within 100 msec, demonstrating real-time responsiveness of the system.

**Extended Data Figure 4.**
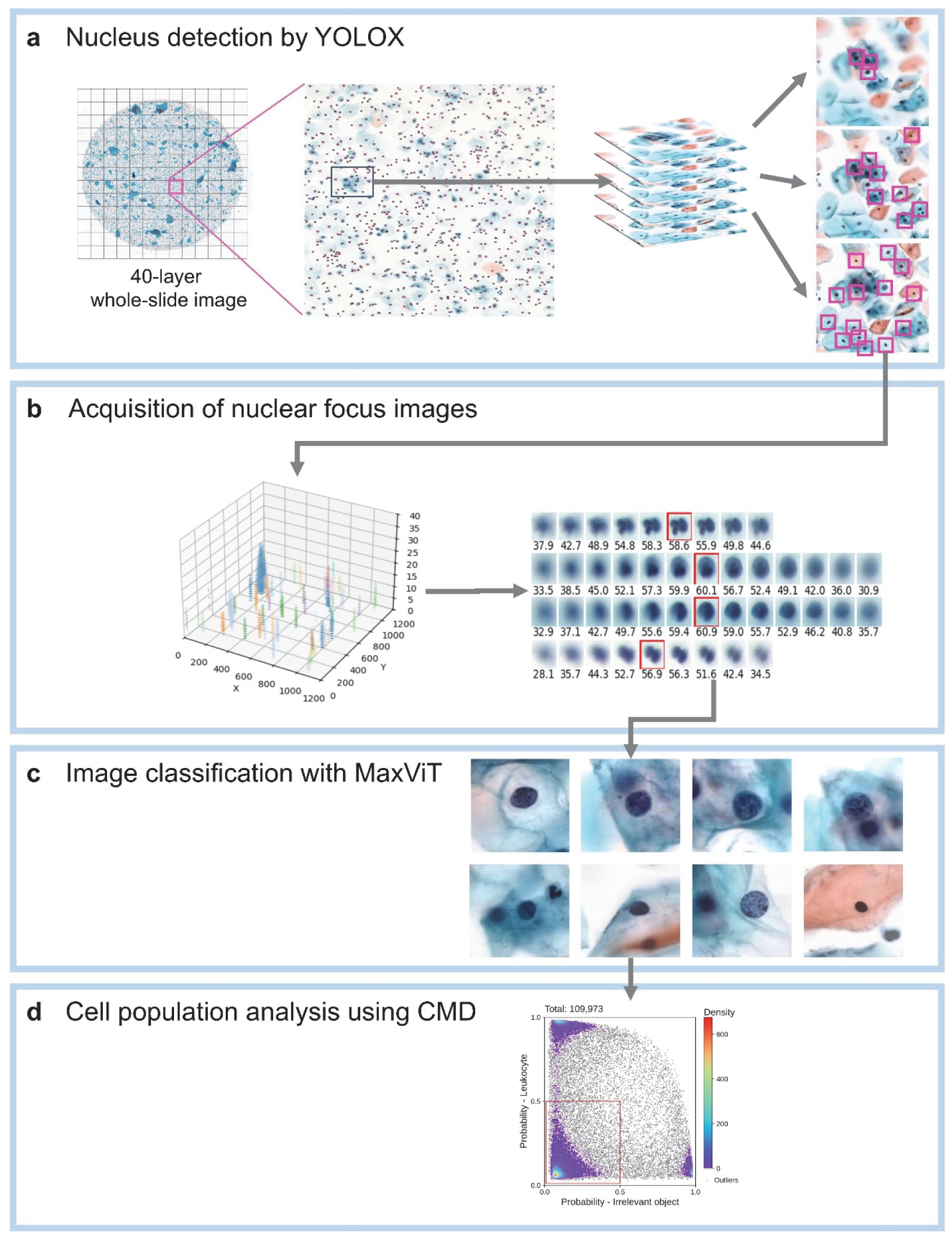
Workflow for 3D whole-slide image analysis of cervical cytology. **a,** Nucleus detection using YOLOX. A 2D object detection model (YOLOX) is applied to sub-sampled optical sections from the 3D Z-stack to identify nuclei. Due to the thickness of the samples, a single nucleus may appear in multiple adjacent layers and thus be detected redundantly. **b,** Acquisition of in-focus nuclear images. Redundant detections are grouped across Z-planes to define individual 3D nucleus instances (left). For each nucleus, a Z-stack of high-resolution patches is retrieved, and the most in-focus slice is selected based on a focus metric (right). **c,** Cell classification with MaxViT. A 224 × 224 pixel patch centered on the nucleus is extracted to include the full cell body. This patch is classified using a MaxViT-based vision transformer model, which outputs a CMD vector – a 10D probability vector representing the cell’s morphology across multiple cytological classes. **d,** Cell population analysis using CMD. CMD vectors from all detected cells are aggregated to enable population-level analysis. This allows for visualization and identification of morphologically similar or diagnostically significant cell clusters across the whole slide.

**Extended Data Figure 5.**
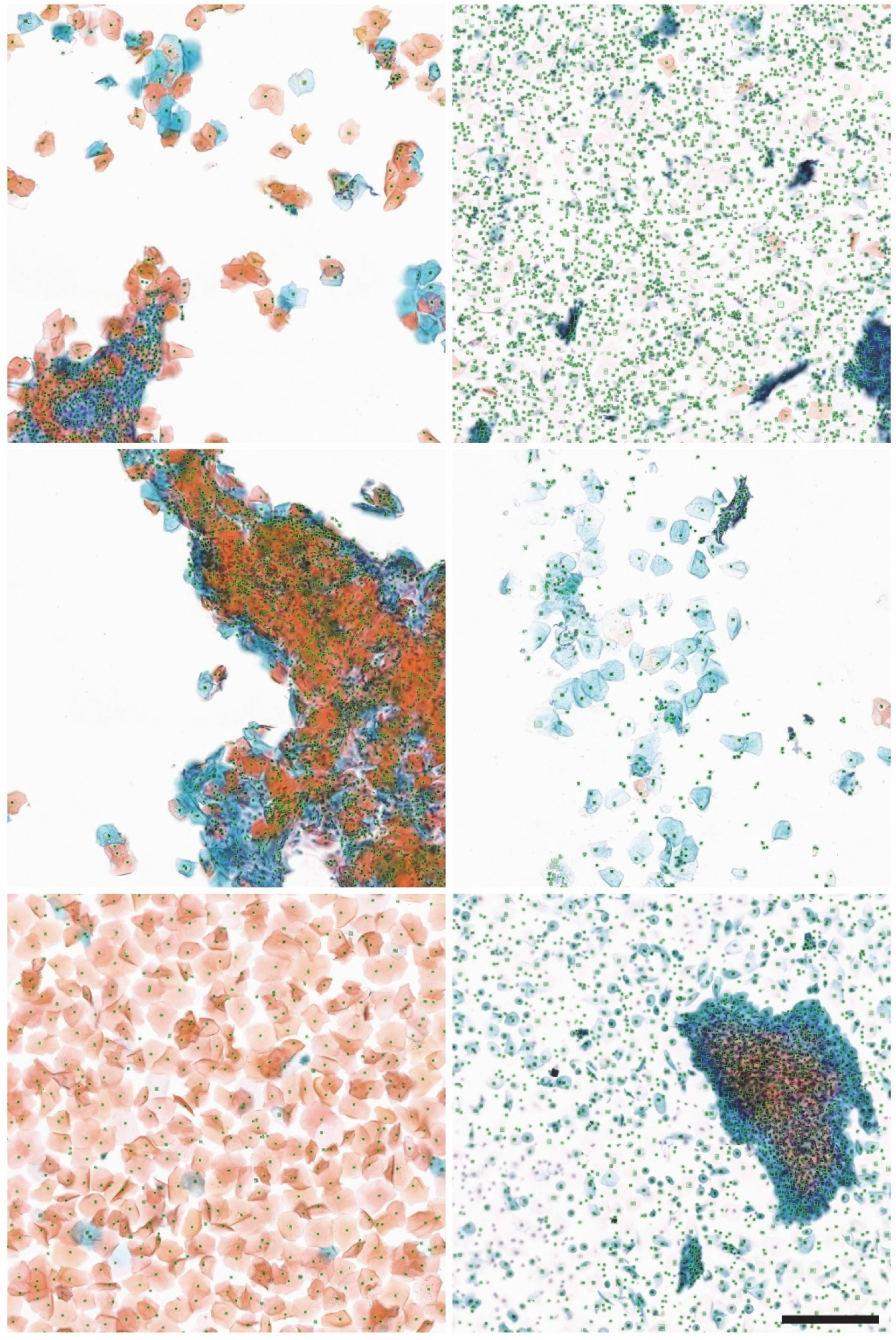
Representative training images used for YOLOX-based nucleus detection. Six representative examples from the training dataset used to develop the YOLOX object detection model. The selected images reflect a wide range of cytological diversity, including variations in cell type, density, and spatial arrangement, to ensure robust nucleus detection across heterogeneous sample conditions. Green bounding boxes indicate manually annotated nuclei that served as ground truth for supervised training. Scale bar: 200 µm.

**Extended Data Figure 6.**
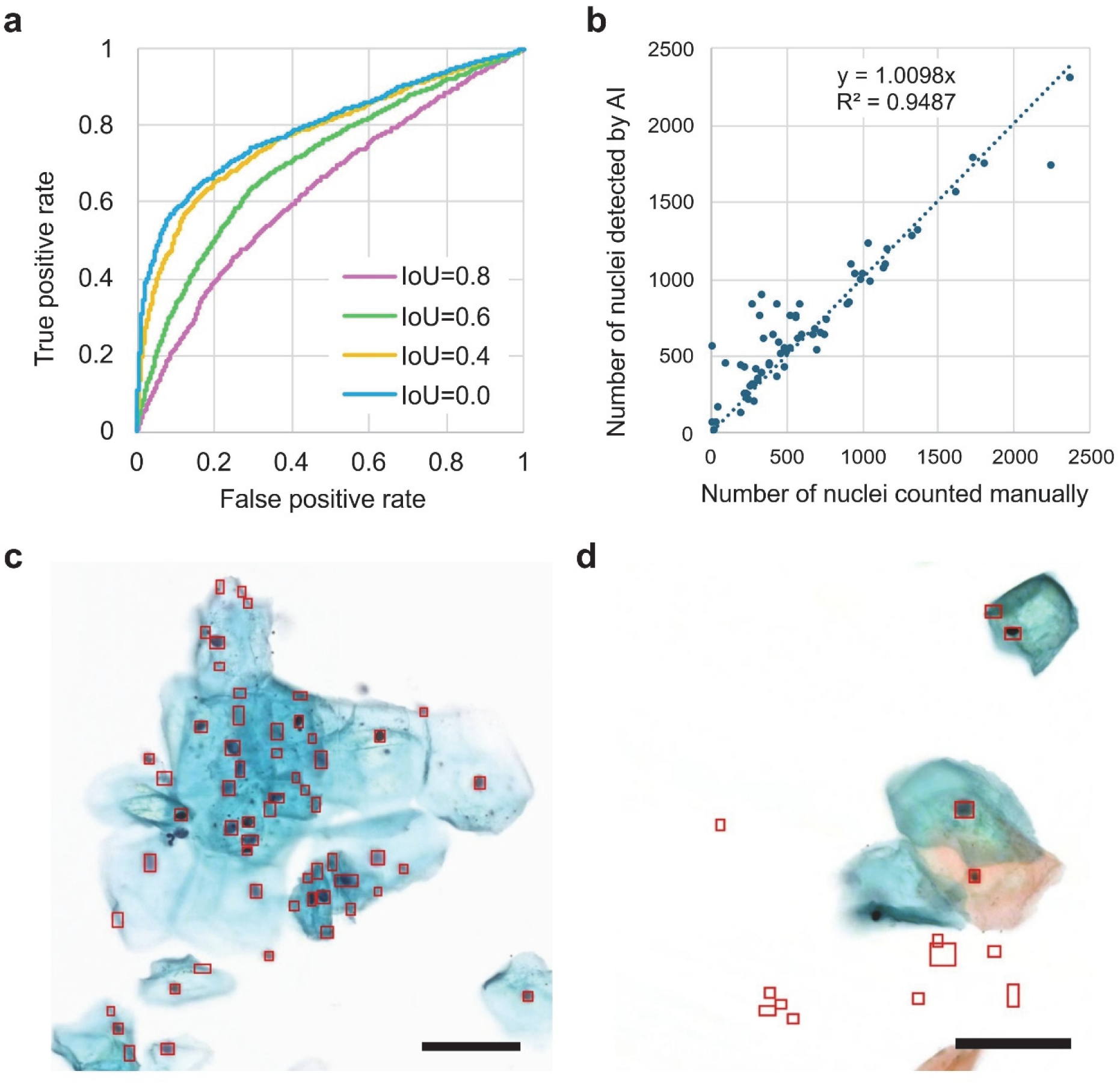
Evaluation of YOLOX-based cell nucleus detection. **a**, ROC curves computed at four IoU thresholds: 0.8, 0.6, 0.4, and 0.0. Model sensitivity and specificity improve as the IoU threshold is relaxed, with the highest AUC observed at IoU = 0.0. **b,** Correlation between manually counted nuclei (x-axis) and YOLOX-detected nuclei (y-axis) across the validation dataset. Each dot represents one image. The regression line and coefficient of determination (R²) indicate strong agreement. **c, d,** Representative outlier cases from panel b in which YOLOX overestimated the number of nuclei. Red bounding boxes highlight all detections, including false positives. Overestimations typically occurred at the periphery of dense cell clusters (**c**) or within background regions lacking cellular content (**d**). Scale bars: 50 µm.

**Extended Data Figure 7.**
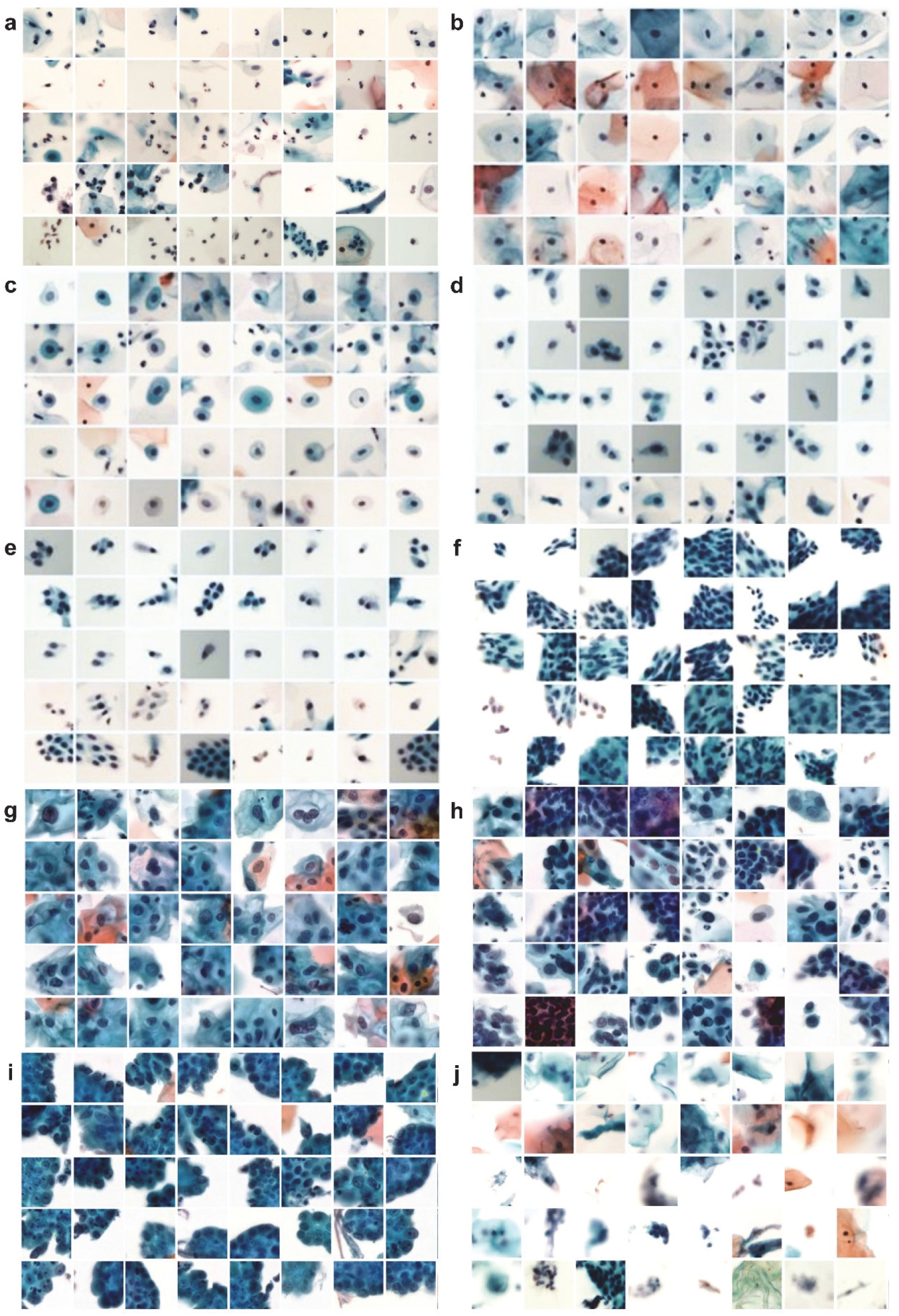
Representative training and validation images used for MaxViT-based cell classification. **a,** Leukocytes. **b,** Superficial/intermediate squamous cells. **c,** Parabasal cells. **d,** Squamous metaplasia cells. **e**, Glandular cells. **f**, Miscellaneous cell clusters. **g,** LSIL cells. **h,** HSIL cells. **i,** Adenocarcinoma cells. **j,** Irrelevant objects (e.g., debris, non-cellular structures, or defocused images). The complete dataset consisted of 18,219 and 5,281 leukocytes, 23,158 and 7,557 superficial/intermediate squamous cells, 4,296 and 1,243 parabasal cells, 2,056 and 487 squamous metaplasia cells, 836 and 105 glandular cells, 5,387 and 994 miscellaneous cell clusters, 1,846 and 936 LSIL cells, 1,433 and 262 HSIL cells, 912 and 420 adenocarcinoma cells, and 14,752 and 5,115 irrelevant objects for training and validation, respectively. Only a small subset of images is shown here for illustration.

**Supplementary Video 1 | Keratinizing squamous cell carcinoma (slide view).** This video demonstrates the digital slide view of a keratinizing squamous cell carcinoma. Scattered tumor cells exhibiting partial keratinization (indicated by orange coloration) are observed against a background of inflammatory cells. The viewer zooms into a representative area of interest, followed by axial navigation through Z-stack layers.

**Supplementary Video 2 | Keratinizing squamous cell carcinoma (tomographic view).** This video presents the tomographic view of the same region shown in Supplementary Video 1. Spindle-shaped cytoplasmic structures of keratinizing squamous cell carcinoma cells are clearly depicted in three dimensions. Although the sample is relatively thin (less than 10 µm), the structural details are well visualized

**Supplementary Video 3 | Non-keratinizing squamous cell carcinoma (slide view).** This video presents the slide view of a non-keratinizing squamous cell carcinoma. A densely packed cluster of tumor cells without keratinization is observed. The video includes zooming into a focal area and sequential Z-stack navigation.

**Supplementary Video 4 | Non-keratinizing squamous cell carcinoma (tomographic view).** This tomographic rendering corresponds to the region displayed in Supplementary Video 3. It highlights a multilayered, sheet-like or solid structure formed by non-keratinizing squamous cell carcinoma cells. The tumor mass demonstrates considerable cell overlapping, with a sample thickness of approximately 20 µm.

**Supplementary Video 5 | HPV-associated adenocarcinoma (slide view).** This video shows the slide view of an HPV-associated adenocarcinoma (usual-type endocervical adenocarcinoma). Tumor cells with a high N/C ratio and finely granular chromatin form a dense cluster, partially exhibiting tubular structures. The video navigates through a representative area of Z-stacks.

**Supplementary Video 6 | HPV-associated adenocarcinoma (tomographic view).** This tomographic view corresponds to the same region shown in Supplementary Video 5. The 3D reconstruction highlights the tubular architecture more clearly, demonstrating the typical morphology of HPV-associated adenocarcinoma. The sample thickness is approximately 20 µm.

**Supplementary Video 7 | HPV-independent adenocarcinoma (slide view).** This video depicts the slide view of an HPV-independent adenocarcinoma, classified as gastric-type mucinous adenocarcinoma. The tumor forms clusters with partially tubular architecture and features abundant mucinous cytoplasm and enlarged round nuclei.

**Supplementary Video 8 | HPV-independent adenocarcinoma (tomographic view).** This tomographic rendering visualizes the same region shown in Supplementary Video 7. It reveals the glandular morphology in three dimensions, with the mucin content within the cytoplasm appearing slightly yellow. The sample represents a gastric-type HPV-independent adenocarcinoma, with a thickness of approximately 15–20 µm.

